# Disease modeling and pharmacological rescue of autosomal dominant Retinitis Pigmentosa associated with *RHO* copy number variation

**DOI:** 10.1101/2023.02.27.23286248

**Authors:** Sangeetha Kandoi, Cassandra Martinez, Kevin Xu Chen, L Vinod K. Reddy, Miika Mehine, Brian C. Mansfield, Jacque L. Duncan, Deepak A. Lamba

## Abstract

Retinitis pigmentosa (RP), a heterogenous group of inherited retinal disorder causes slow progressive vision loss with no effective treatments available. Mutations in the rhodopsin gene (*RHO*), account for ∼25% cases of autosomal dominant RP (adRP). In this study, we describe the disease characteristics of the first ever reported mono-allelic copy number variation (CNV) in *RHO* as a novel cause of adRP. We (1) show advanced retinal degeneration in a male patient (60-70 year old) harboring four transcriptionally active intact copies of rhodopsin, (2) recapitulated the clinical phenotypes using retinal organoids, and (3) assessed the utilization of a small molecule, Photoregulin3 (PR3), as a clinically viable strategy to target and modify disease progression in RP patients associated with *RHO*-CNV. Patient retinal organoids showed photoreceptors dysgenesis, with rod photoreceptors displaying stunted outer segments with occasional elongated cilia-like projections (microscopy); increased *RHO* mRNA expression (qRT-PCR and bulk RNA-sequencing); and elevated levels and mislocalization of rhodopsin protein (RHO) within the cell body of rod photoreceptors (western blotting and immunohistochemistry) over the extended (300-days) culture time period when compared against control organoids. Lastly, we utilized PR3 to target *NR2E3*, an upstream regulator of *RHO*, to alter *RHO* expression and observed a partial rescue of RHO protein localization from the cell body to the inner/outer segments of rod photoreceptors in patient organoids. These results provide a proof-of-principle for personalized medicine and suggest that *RHO* expression requires precise control. Taken together, this study supports the clinical data indicating that adRP due to *RHO*-CNV develops due protein overexpression overloading the photoreceptor post-translational modification machinery.

## Introduction

Retinitis pigmentosa (RP) is a genetically heterogenous group of ‘rod-cone’ photoreceptor degenerative diseases that are unified by common clinical features characterized by progressive vision loss, commonly starting as night blindness(O’Neal and Luther 2023). RP affects roughly 1 in 3000-5000 individuals and can be inherited as autosomal recessive, autosomal dominant (ad) or X-linked disease(Chizzolini et al. 2011). adRP can be caused by mutations in at least 24 different known genes (“RetNet: Summaries” n.d.)amongst which mutations in rhodopsin gene (*RHO*) accounts for 25% of total adRP cases(Meng, Ragi, and Tsang 2020). A mutation in the *RHO* gene was the first identified cause of RP due to a single-base substitution at codon 23 (P23H) leading to protein misfolding and triggering the death of the rod photoreceptors(Dryja et al. 1990). *RHO* is located on the long arm of chromosome 3 (3q22.1) and drives the expression of a 348 amino-acid G protein-coupled receptor (GPCR) with seven transmembrane domains, a luminal N terminus, and a cytoplasmic C terminus. Rhodopsin protein (RHO) is localized in the densely packed disc membrane of the rod photoreceptor outer segments. Currently >150 different rhodopsin mutations have been identified, all contributing through multiple mechanisms with each having distinct consequences on the protein structure and function(Athanasiou et al. 2018). Based on the experimentally studied biochemical and cellular characteristics, several mechanisms have been linked with *RHO*-mutation to cause photoreceptor degenerations including protein misfolding, retention and instability in endoplasmic reticulum (ER), glycosylation defects, post Golgi trafficking and outer segment targeting, dimerization deficiency, altered post-translational modifications and reduced stability, disrupted vesicular trafficking and endocytosis, and impaired trafficking, leading to constitutive phototransduction activation or altered transducin interactions(Newton and Megaw 2020).

In a conference proceedings, we reported copy number variations (CNV) in *RHO* as a novel cause of adRP(Duncan et al. 2019). The current study presents a unique opportunity to better understand the pathogenic effects of two extra copies of intact wild-type *RHO* on a single allele at 3q22.1 in a male patient (60-70 year old) diagnosed with adRP. Transgenic mice overexpressing wild-type *Rho* have previously demonstrated photoreceptor degeneration, however the precise mechanism of degeneration is still unclear(Olsson et al. 1992). Although mice have similar genetics to humans, the distribution, subtypes, quantity of retinal cells (especially photoreceptor cells), and the developmental timeline of the retina differs greatly(Volland et al. 2015). Therefore, access to human cells and tissues *vis-à-vis* the retinal organoid model provides a reliable, translational, and clinically relevant system to gain insights in understanding the pathogenic effects of excessive rhodopsin in photoreceptors. Induced pluripotent stem cell (iPSC)-based models have been used to model several retinal degenerations such as Retinitis Pigmentosa (Tucker et al. 2013; Giacalone et al. 2019), Usher’s syndrome (Dulla et al. 2021), Leber congenital amaurosis (LCA) (Parfitt et al. 2016; Kruczek et al. 2021) and *CRX*-associated LCA7 (Chirco et al. 2021). This report aimed at presenting the late-onset *RHO*-CNV associated with Retinitis Pigmentosa expands the spotlight on modeling a novel cause of retinal disease *via* a 3D ‘disease-in-a-dish’ platform.

Patient-specific retinal organoids serves as a versatile tool for testing various therapeutic interventions including small molecule drugs which aim at modulating the pathways (Moore, SkowronskaLKrawczyk, and Chao 2020; Liu et al. 2021). Small molecule-based targeted therapeutics have the potential to cross the blood-brain barrier when administered systemically and can be therapeutically titrated. Amongst these, nuclear receptors are important targets as they have druggable ligand-binding sites. Approximately 15% of approved drugs, target at least 48 members of human nuclear receptors superfamily, and 10 amongst these are orphan(Zhao, Zhou, and Gustafsson 2019). Numerous orphan nuclear receptors are expressed in the retina, of which rod-specific nuclear receptor subfamily 2 group E member 3 (*NR2E3*) is a direct target of neural retina leucine zipper (*NRL*), the main rod-specifying gene(Kobayashi et al. 1999). *NR2E3* is expressed very early in post-mitotic rods and coactivates the transcription of rod-specific genes including *RHO*, with *CRX* and *NRL*(Bumsted O’Brien et al. 2004; Cheng et al. 2004; Mitton et al. 2000) while suppressing cone genes. Recent studies have identified photoregulins (PR), small molecules that can target NR2E3. These molecules, especially PR3, have been demonstrated to reduce expression of rhodopsin and other rod genes and modify disease progression in mouse models of rod photoreceptor mutation-associated RP (Nakamura et al. 2016, 2017). We tested the hypothesis that PR3 can mitigate the deleterious effects of the *RHO*-CNVs on *in vitro* patient-specific retinal organoids by reducing expression of RHO and allowing normal cellular processing. Overall, we demonstrated the establishment of a human retinal organoid model of *RHO*-CNV associated with adRP to gain critical insights into the pathogenesis of disease and to provide a vital screening tool for the development of various novel therapies.

## Results

### *RHO*-CNV patient presented clinical features of adRP

The clinical and ERG data of one patient and one healthy unaffected daughter of the patient is included (**Table 1**). Complete ophthalmological examination by fundus photography and SD-OCT revealed features of RP (Hirji 2023) including bone spicule-like pigmentation changes, optic disc pallor and attenuation of retinal blood vessels (**Figure 1A, Supplementary Figure 1**), with outer retinal atrophy due to the loss of the photoreceptor layers, sparing the central foveal region (**Figure 1B**). While the central foveal region was relatively spared in this patient, the macular cones that remained were observed to be damaged by chronic edema, and photoreceptor and RPE atrophy had progressed into the macula. Genetic testing of the proband by next-generation sequencing (NGS) showed a complex duplication-rearrangement of chromosome 3q22, which encompassed the entire *RHO* coding sequence, 5’ and 3’ regulatory regions and flanking genes. The rearrangement consisted of a 48 kb triplicated region embedded within a 188 kb duplication, resulting in three apparently intact *RHO* genes on one chromosome and a fourth, unaltered *RHO* gene on the homologous chromosome. Within the duplications, *RHO* copies were flanked by *H1FOO*, *IFT122*, *EFCAB12* and *MBD4* while the inverted triplication of *RHO* was flanked by partially triplicated *IFT122* and *H1FOO* (**Figure 1C**). Thus, the NGS data shows a duplication-inverted triplication-duplication event, in which the triplicated segment is inverted and located between correctly oriented genomic segments (**Figure 1C**). No other additional causative variants of inherited retinal degeneration candidate genes were identified. Whole genome sequencing supported the rearrangement and extra copies of *RHO* which were identified by NGS. The *RHO* copy number variants were not detected in the unaffected daughter of the patient. Clinically collected four-generation pedigree data of the family included two male family members affected with RP, indicating male-male transmission, consistent with an autosomal dominant inheritance. The proband’s affected father was not examined as he was deceased but had been diagnosed clinically with RP. The clinical findings were consistent with the hypothesis that CNV in the wild-type *RHO* causes adRP.

**Figure 1:**
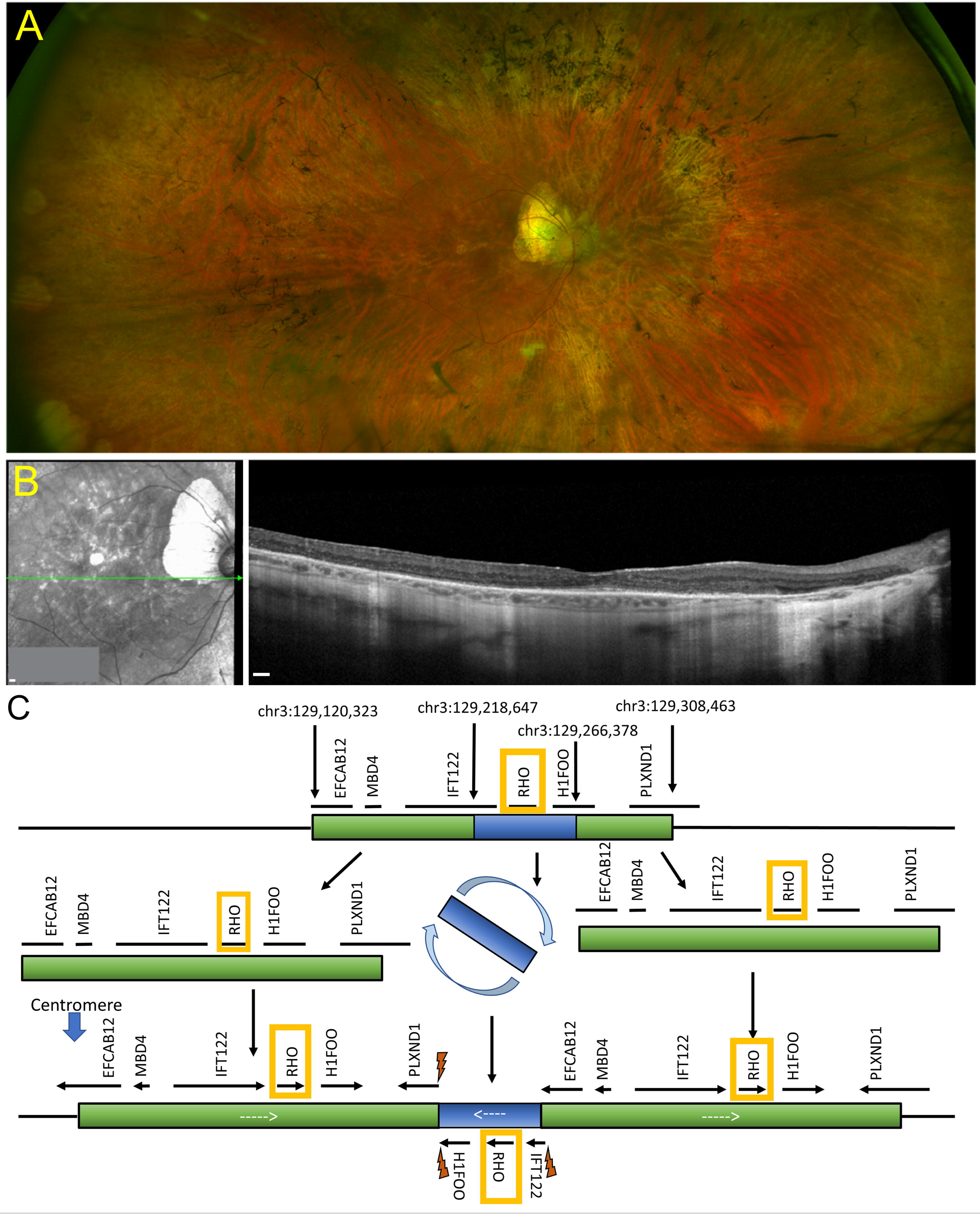
Retinal imaging and next generation sequencing. **(A)** Ophthalmological color fundus examination of a patient clinically diagnosed of *RHO-*CNV displaying bone spicule pigmentary changes. **(B)** SD-OCT image showing extensive loss of the outer retinal layers leaving a small intact island at the fovea. **(C)** Schematic illustration of NGS using a 266-gene retinal dystrophy panel showing a complex chromosome 3q22 duplication-rearrangement resulting in an inverted 48 kb triplicated region embedded within a 188 kb duplication forming three apparently intact *RHO* genes on one allele and a fourth, unaltered *RHO* on the homologous allele. Lightning bolts represent genomic breakpoints of triplication insertion. *RHO* was the only gene that is fully triplicated. NGS=Next generation sequencing; SD-OCT= Spectral domain-optical coherence tomography. Scale bar = 200µm in A and B.

### *RHO*-CNV retinal organoids exhibit photoreceptor maturation defects

To assess the effects of *RHO*-CNV in human retinal organoids, we initially reprogrammed the peripheral blood mononuclear cells (PBMNCs) from one patient with four copies of *RHO* (RM) as well as the corresponding familial control (RC) into iPSCs (**Supplementary Figure 2A-B**). The iPSC lines had the colony morphology of tightly packed cells with a high nucleus-to-cytoplasm ratio, a well-defined border typical of stem cells, and expression of pluripotent markers (**Supplementary Figure 2C-D**). We then differentiated the patient and control iPSC lines into retinal organoids by following our previously published protocols(Bachu et al. 2022; Arthur et al. 2022), as per the differentiation timeline depicted in **Figure 2A**. Retinal organoids from the *RHO*-CNV patient and the control displayed a well-defined neuroepithelial lamination, indicating the arrangement of photoreceptors on the apical surface of the organoids and a dark central basal region consisting of inner retinal cells by phase contrast microscopy (**Supplementary Figure 2E**). Notably, there were no clear visible anatomical changes in apical-basal retinal cell type distribution during the early differentiation timeframe, when the cone and rod photoreceptors are usually born in 45-50-days-old and 90-120-days-old human retinal organoids, respectively (data not shown). Over the prolonged differentiation culture timeframe (>200 days), the control retinal organoids displayed long hair-like protrusions which were presumptive inner and outer segments at the apical side of the retinal organoids, a critical event indicating the start of photoreceptor maturation. Conversely, the patient retinal organoids showed short initial hair-like protrusions that did not elongate at the extended culture time of 260 and 300 days in culture by light microscopy (**Supplementary Figure 2E**, **Figure 2B**). To validate these observations, we examined the ultrastructure of photoreceptors in the *RHO*-CNV and control organoids using transmission electron microscopy (**Figure 2C, Supplementary Figure 3E**). Upon assessing the 300-days-old organoids, we observed that while the patient organoids developed connecting cilium and inner segments similar to control organoids, they failed to extend outer segments. This data was consistent across multiple rounds of differentiation from three independent clones of patient iPSCs. Taken together, these morphological changes suggest that iPSC-retinal organoids of *RHO*-CNV patient demonstrated the survival of photoreceptors with OS dysmorphogenesis over the time course of 300-days.

**Figure 2:**
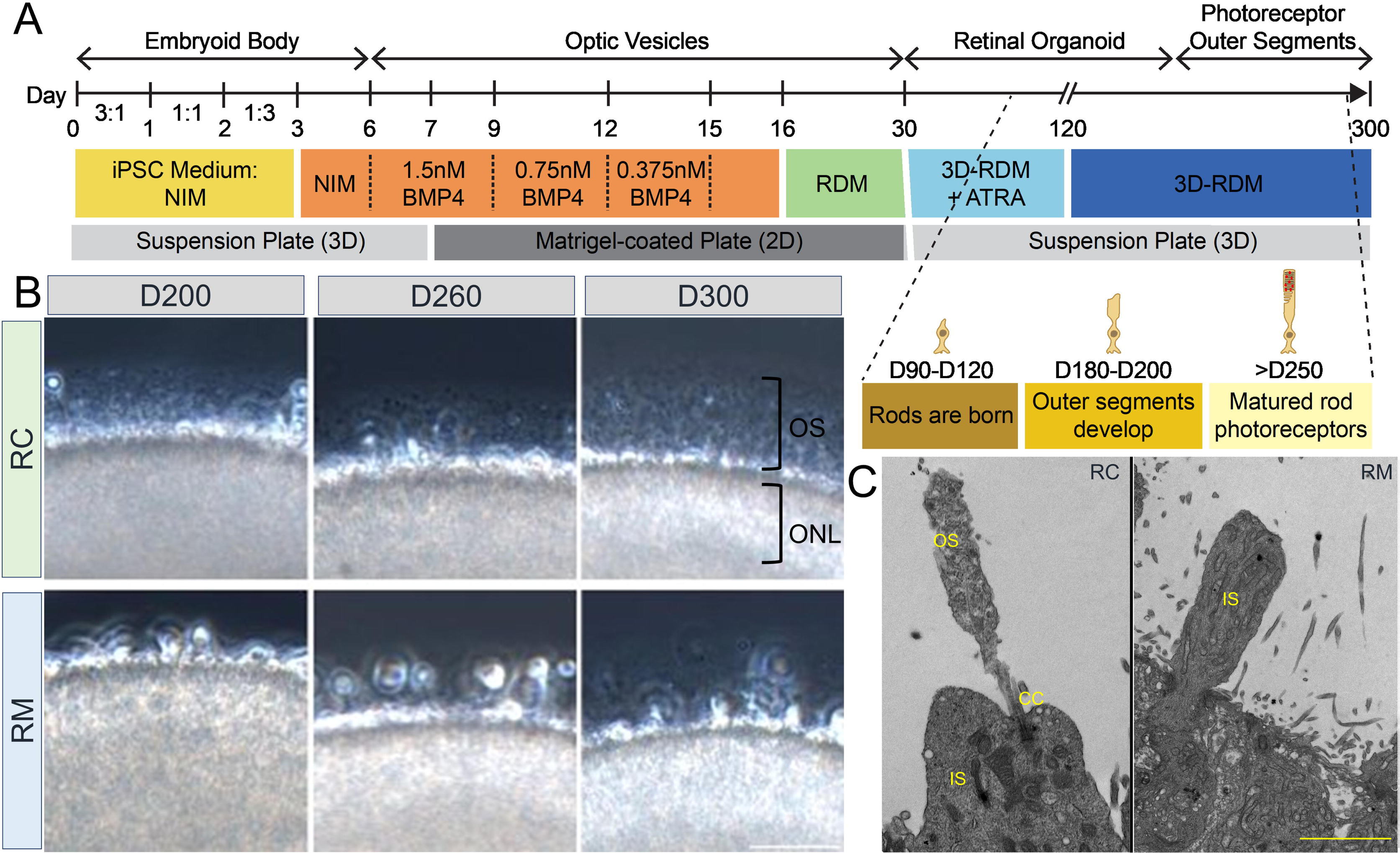
*RHO*-CNV disease modeling using iPSC-derived retinal organoids showed morphological defects. **(A)** Schematic representation showing the timeline of human retinal differentiation and maturation including the birth and development of rod photoreceptors. **(B)** Phase contrast microscopy images showing OS, long hair-like protrusions from ONL of the differentiated photoreceptors present at the apical surface of control (RC) retinal organoids at day 200, 260 and 300 (top). Conversely, retinal organoids from patient (RM) showing shorter protrusions which do not extend progressively over long-term culturing indicating maturation defects (bottom). **(C)** Electron microscopy images showing ultra-magnification of distinct OS, IS and CC structures of rod photoreceptors in control organoids and the absence of OS in patient organoids. CC=Connecting cilium; IS=Inner segments; ONL= outer nuclear layer; OS=Outer segments. Scale bar = 50 µm.

### *RHO*-CNV retinal organoids revealed conspicuous defects in rod phototransduction and ciliary transcripts

Control and patient retinal organoids were analyzed at two developmental stages: at rod photoreceptor birth (D120), and maturation (D300). For each sample, qRT-PCR was carried out by utilizing primers designed to nine key genes that regulate the development and maturation of rod photoreceptors. mRNA levels were analyzed for the expression of genes specific to early pan photoreceptors (*OTX2, CRX, RCVRN*), early rod photoreceptors (*NRL, NR2E3*), rod-specific phototransduction (*PDE6B, SAG, RHO*) and ciliary (*IFT122*) genes (**Supplementary Table 4**). To equilibrate the data to equivalent number of photoreceptors in organoids, we normalized the data to *CRX* expression, a transcription factor highly enriched in photoreceptors in the retina (Yamamoto et al. 2020). The expression of all the target genes were detected at each time point (D120 and D300) in the retinal organoids (**Figure 3A**). Pan-photoreceptor and early rod marker genes showed similar expression levels with no noticeable variations in RC and RM organoids. In contrast, RHO and SAG were expressed at a higher level in the patient compared to control organoids. There was a significant eight fold increase in the *RHO* levels at D120 and D300 in the patient organoids compared to controls. We also observed a small two-fold statistically significant increase in rod/visual arrestin (*SAG*) at D300 timepoint in the patient organoids compared to control organoids. Relatedly, a one-fold, but non-statistically significant, increases in patient organoids was observed in *IFT122, a* gene partially triplicated in NGS along with *RHO* (**Figure 1C**).

**Figure 3:**
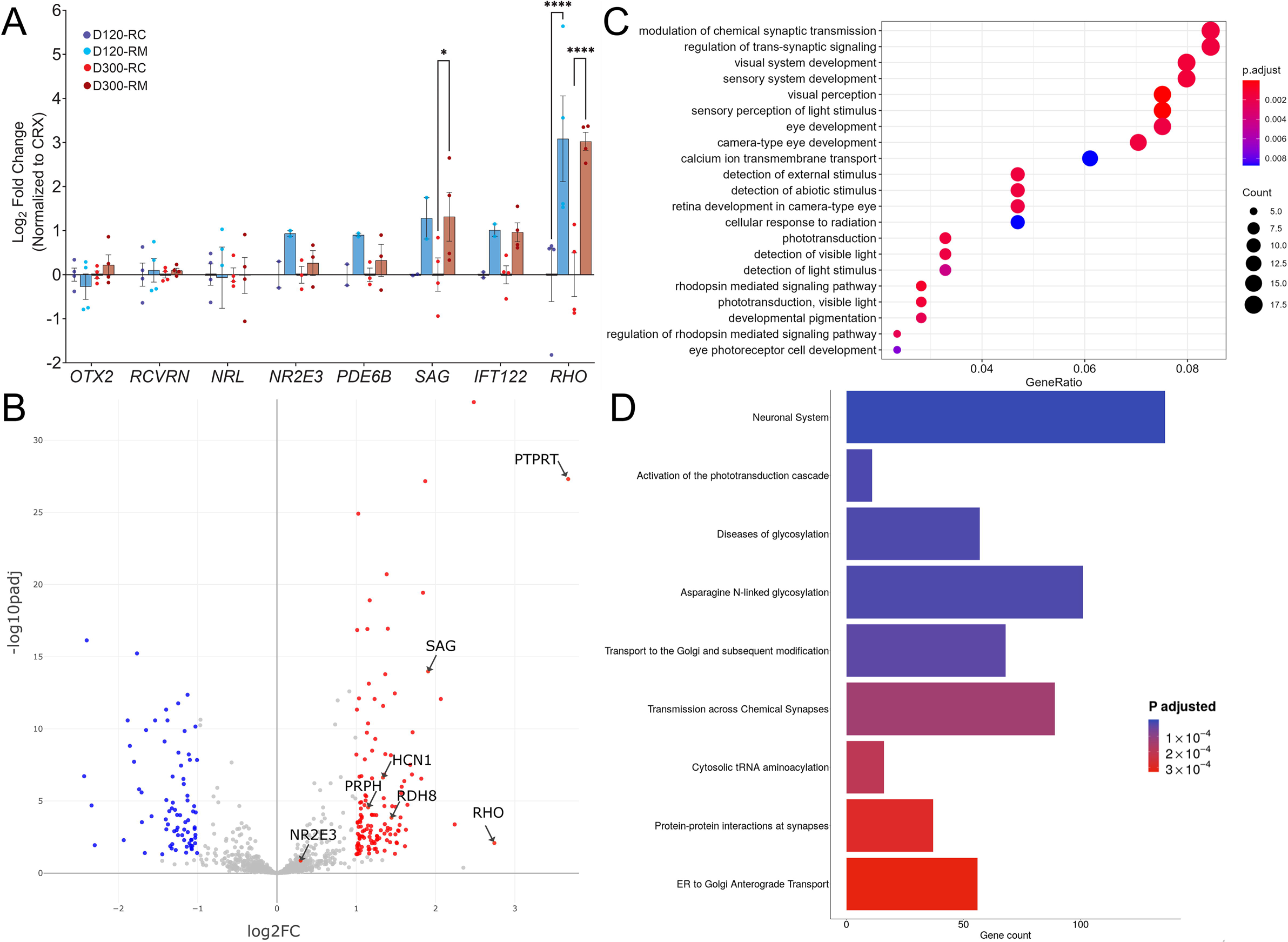
Transcriptomic analysis of *RHO*-CNV retinal organoids presented elevated RHO expression. **(A)** qRT-PCR analysis shows eight fold increase in *RHO* mRNA levels in patient (RM) organoids compared to control (RC) at D120 and D300 of rod differentiation and maturation. No significant change was observed in other photoreceptor genes except for a small two-fold increase in rod arrestin (SAG) at D300 time-point. Log2FC=Log 2 Foldchange. Statistical two-way ANOVA analysis with Fisher’s LSD test and 95% confidence interval. * = p <0.05, and **** = p <0.0001. Blue bars, D120; Maroon bars, D300 **(B)** Volcano plot showing significant differentially expressed genes following bulk RNAseq analysis comparing patient to control organoids (>D300). Significantly upregulated genes are highlighted in red and significantly downregulated genes are highlighted in blue (adjusted p<0.01). **(C)** Dot plot showing EnrichGO analysis of biological process on the differentially expressed genes in the bulk RNAseq analysis. The size of the dot represents number of differentially expressed genes in the pathway and the X-axis represent the ratio over all genes associated with the pathway. Plot shows a defect in rods and phototransduction associated pathways as well as synaptic transmission suggesting rod dysfunction. **(D)** Box plot showing the data from pathway enrichment analysis of cellular component category predominantly highlighting the defect in glycosylation and Golgi/ER modification/transport. Colors in the dot and blot plots represent relative significance (calculated p-values in scale). N=3-4 independent experiments and 12-15 organoids per experiment.

To advance a better molecular understanding on detailed genomic expression and gene regulatory networks, we performed bulk-RNA sequencing comparing 300-days-old retinal organoids from patient and control iPSCs (n=3 independent biological replicates). Patient retinal organoids demonstrated upregulated transcriptomic levels of *RHO* (∼eight-fold) and *SAG* (∼four-fold) compared to control organoids, comparable to the differences observed in the qRT-PCR data (**Figure 3B**). Additionally, we also observed increased expression in disc structure support gene including *PRPH*, visual cycle genes *RDH8* and *HCN1*, and a synaptic gene, *PTPRT* in patient relative to control organoids (**Figure 3B**). Following the Gene Ontology enrichment analysis (EnrichGO) of significant differentially up-regulated genes, we confirmed increases in phototransduction cascades including the visual/light perception and membrane potential/Ca+ signaling pathways in patient organoids compared to control organoids. Additionally, increases in genes associated with the synaptic signaling pathway were also detected, suggestive of photoreceptor dysfunction (**Figure 3C**). Pathway enrichment analysis of significantly differentially expressed genes for cellular components category pointed to perturbation in pathways associated with defects in glycosylation especially N-linked glycosylation as well as ER and Golgi transport (**Figure 3D, Table 2**), highlighting potential pathophysiology that may drive *RHO*-CNV associated RP. Although glycosylation is not required for the rhodopsin biosynthesis, N-linked glycosylation (at N2 and N15), a post-translational modification, is a necessary step for interacting with chaperones during ER transport. This is also an essential step towards incorporation of the heptahelical G-protein coupled receptor rhodopsin in the rod outer segments. Thus, RNAseq data suggests that the defects in rhodopsin glycosylation decreased the ability of rhodopsin to exit ER, and lead to an adRP phenotype (Tam and Moritz 2009; C.LH. Sung and Tai 1999).

### *RHO*-CNV retinal organoids displayed mis-localized and elevated Rhodopsin protein levels

Immunofluorescence staining on the cross sections of control and patient retinal organoids was examined for spatial location of the photoreceptors at three time points (D120, D200, D300). Pan-photoreceptor precursor and progenitor proteins (OTX2, CRX), and rod specific proteins (NR2E3) were expressed in the apical layer of the organoids in a similar pattern both in the control and patient organoids with equivalent photoreceptors and outer nuclear layer thickness (**Supplementary Figure 3A-C**). These results corroborated our gene expression and transcriptomics data (**Figure 3A-B**) indicating no defects in rod biogenesis within the patient retinal organoids. As the organoids began to mature, the distribution of RHO protein was observed in the outer segments of control organoids starting at D200, earliest timepoint for detection in human retinal organoids using our differentiation protocol. Compared to the control, the patient organoids had mis-localized RHO protein accumulating in the photoreceptor cell soma, at all analyzed time-points (D200, and >D250) (**Figure 4A-B**). The patient organoids rarely showed RHO in IS/OS which were notably consistent with our results discerned by light microcopy (**Supplementary Figure 2E**, **Figure 2B**). Co-staining with NRL and CRX confirmed mislocalized RHO expression in the rod photoreceptor soma within patient organoids. We also evaluated the expression of rod-specific phototransduction protein, SAG, which was increased in the outer nuclear layer of patient retinal organoids (**Figure 4C**). We did not observe any differences in the cone markers, ARR3, BCO or pan phototransduction marker, RCVRN when comparing >D250 patient to control organoids (**Supplementary Figure 3D**). There was also no difference in apoptosis between the patient and control organoids (**Supplementary Figure 3F**).

**Figure 4:**
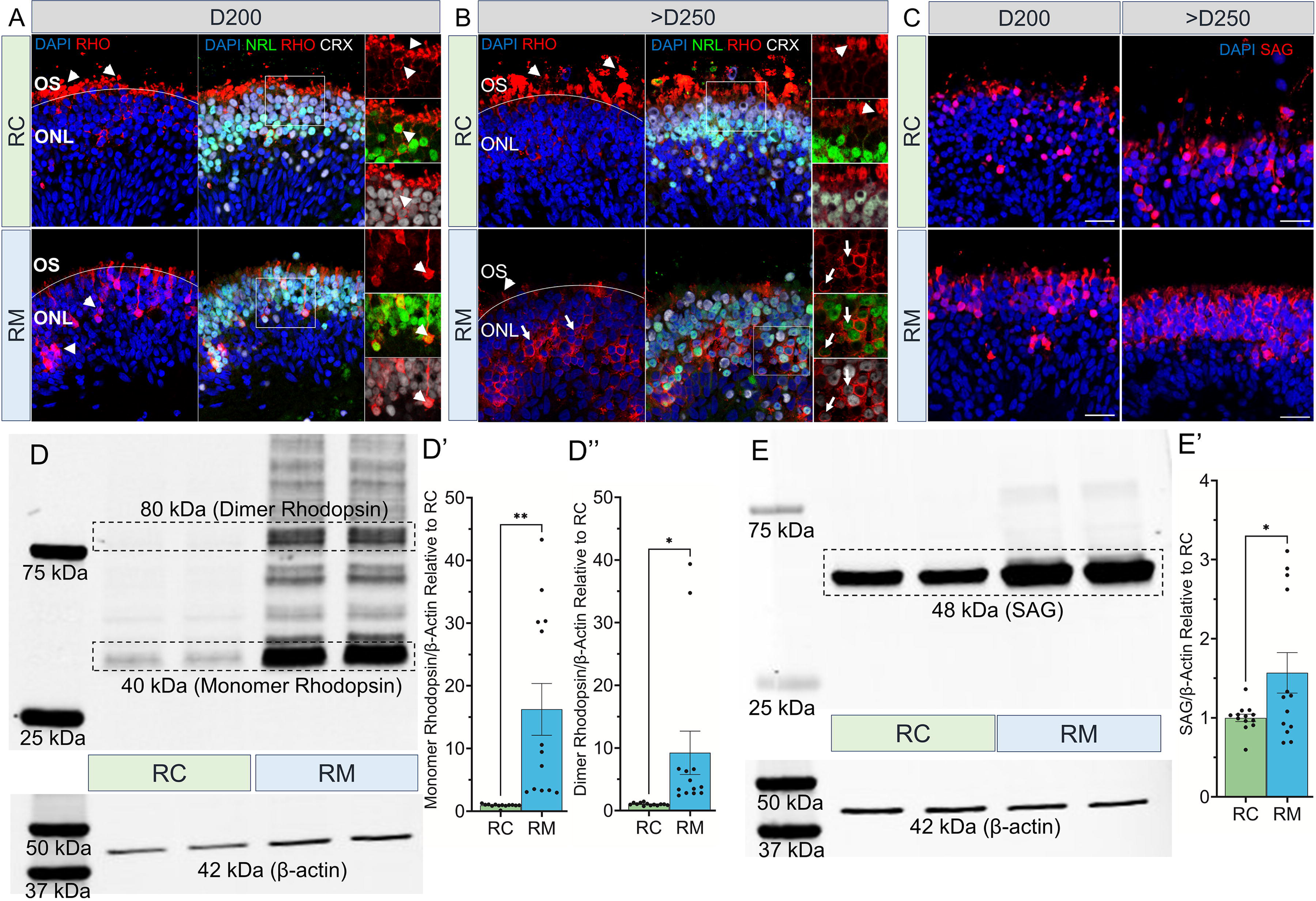
Rhodopsin protein mislocalization and increased levels in *RHO*-CNV retinal organoids. **(A-B)** Immunofluorescence staining of RHO, NRL and CRX displaying the proper RHO localization (arrowheads) in the outer segments of control (RC, top panel) organoids and RHO mislocalization (arrows) in the cell body of photoreceptors within the patient (RM; bottom) organoids at two time-points, D200, and >D250. Occasional inner/outer segments with appropriate RHO localization in the patient (RM) organoids were seen at >D250 time-point (arrowhead). (**C)** SAG expression (red) was also increased in the cell soma of patient organoids (bottom) compared to control organoids (top). DAPI (blue) labels nuclei. OS=outer segment; ONL=outer nuclear layer. Scale bar = 25 µm. **(D-E)** Western blot probed for RHO and SAG showing increased levels of 40 kDa monomer and 80 kDa dimer, RHO **(D)**, and 48 kDa, SAG **(E)** in patient retinal organoids compared to controls. β-actin was used as a loading control. Densitometric analysis quantifying the relative intensity of monomeric RHO **(D’)**, dimeric RHO **(D’’)**, and SAG **(E’)** in comparisons to the control. Statistical two-tailed unpaired T-test analysis with 95% confidence level. * = p <0.05, and ** = p <0.01. N=5 independent experiment and 12-15 organoids per experiment.

To investigate protein expression levels in *RHO*-CNV organoids, we quantitated the level of RHO and SAG by western blot by comparing to control organoids. Patient retinal organoid homogenates displayed a significant 16-fold and 9-fold higher fractions of ∼40 kDa monomer and ∼80 kDa dimer rhodopsin content respectively in patient organoids relative to controls despite loading equal amounts of protein lysates by western blot (**Figure 4D-D’’**). A significant 1.5-fold increase in ∼48 kDa SAG, a rhodopsin interacting protein was also observed (**Figure 4E-E’**). Additionally, we analyzed the ER stress-UPR pathway in the patient organoids and did not detect any differences compared to control (*data not shown*). These findings suggest that *RHO*-CNV organoids actively translated the excessive rhodopsin protein which is not getting transported to the outer segments.

### PR3 treatment attenuates *RHO* expression and partially rescues RHO trafficking in *RHO*-CNV retinal organoids

Since, the excess RHO protein production due to the extra *RHO* copies is likely not being processed appropriately, we aimed to test potential therapies that may reduce RHO protein levels in rods. We targeted orphan nuclear receptor, *NR2E3*, using a small-molecule, PR3, to assess its effect on *RHO* regulation and expression in the human patient organoid model (**Figure 5A**). PR3has been previously described to regulate the rod gene expression in Rho^P23H^ mice following a screen for molecules targeting NR2E3, suggesting its potential for the use in the treatment of RP (Nakamura et al. 2017). We treated long-term cultured 300-days-old, *RHO*-CNV patient retinal organoids with varying concentrations of PR3 (0.1, 0.25 and 0.5 µM) for one week and assessed the effects on *RHO* mRNA expression and protein localization. Immunofluorescence staining of PR3-treated organoids displayed a partial rescue of RHO localization (**Figure 5B**). 0.5 µM treatment lead to the strongest reduction in RHO with very few rods still expressing the protein. 0.25 µM treatment lead of best outer segment region localization of RHO. None of the organoids showed any evidence of toxicity (by TUNEL, **Supplementary Figure 4C**) post-treatment. Following qRT-PCR analysis of PR3 treated organoids compared to vehicle treated ones, we observed a four to thirty fold decrease in *RHO* expression in a dose-dependent manner, along with smaller decreases in other rod-specific genes including *NR2E3*, *GNAT1 and PDE6B* (**Figure 5C**). We did not see any significant effects of PR3 on blue-, green- and red-cone opsin genes or protein expression for these protein or cone arrestin (**Supplementary Figure 4A, B**). Upon comparison of PR3-treated patient organoids with control (RC) organoids. *RHO* expression levels in the patient organoids treated with 0.1 and 0.25 µM PR3 was not significantly different from control organoids. However, 0.5 µM PR3 resulted in significantly decreased *RHO* expression, much lower than the *RHO* levels observed in control organoids (**Figure 5D**).

**Figure 5:**
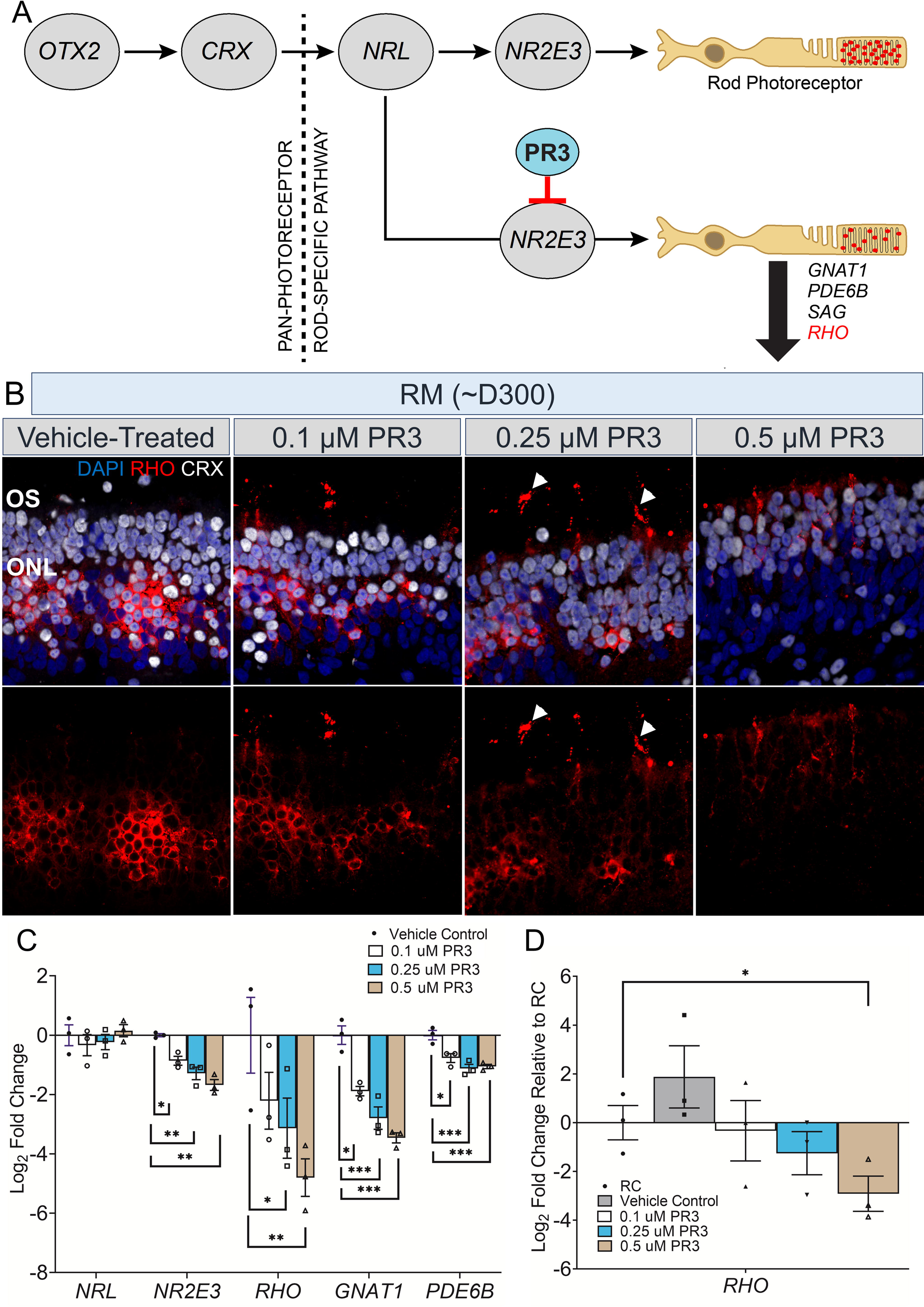
Partial rescue of rhodopsin localization and expression levels in PR3 treated *RHO*-CNV retinal organoids. **(A)** Cartoon illustration showing gene expression during the stepwise development and maturation of rod photoreceptors. Small molecule, PR3 is predicted to act on *NR2E3* downregulating the expression of *GNAT1, PDE6B, SAG* and *RHO*. **(B)** Immunostaining of ∼300-days-old PR3-treated retinal organoid sections from patient (RM) showing the trafficking of RHO protein (arrowheads) towards outer segments at all three doses of PR3, 0.1, 0.25 and 0.5 µM. Retinas co-stained for CRX to mark photoreceptors. The most appropriate RHO localization to OS is seen at 0.25 µM PR3 (arrowheads) and much lower overall RHO expression in 0.5 µM PR3. DAPI (blue) labels nuclei. OS=outer segment; ONL=outer nuclear layer. Scale bar =25 µm. **(C)** qRT-PCR analysis shows eight to 30 fold decrease in *RHO* mRNA levels in a dose-dependent manner for PR3 treated-patient (RM) organoids. No significant change was observed in *NRL*, but a small decrease was observable in *GNAT1, PDE6B* and *NR2E3* (one-way ANOVA analysis with Sidak test and 95% confidence interval). **(D)** qRT-PCR analysis showing a comparison of *RHO* mRNA levels in PR3-treated patient organoids to control (RC) organoids (unpaired t-test). Log2FC=Log2 Fold change. * = p <0.05, ** = p <0.01and *** = p <0.001. N=3-4 independent experiments and 12-15 organoids per experiment.

We further carried out bulk RNA-sequencing analysis to comprehensively characterize three different groups of organoids, 0.25 µM PR3-treated and vehicle-treated patient organoids and control (RC) organoids from three independent differentiation experiments. Consistent with the qRT-PCR gene expression analysis, the results showed a significant downregulation in *RHO* and other rod phototransduction genes including *SAG* and *GNAT1* between the PR3 and vehicle treated patient organoids (**Figure 6A**). Additionally, we confirmed that PR3 did not have any adverse effects on cone opsin transcripts in patient organoids. Principal component analysis (PCA) and normalized read counts analysis of sequenced data for rod (*RHO, SAG, GNAT1*, *NR2E3*) and other genes demonstrated that the PR3-treated organoids were more alike to control organoids than the vehicle treated patient organoids (**Figure 6B-C, Supplementary Figure 5B**). Upon KEGG analysis of differentially expressed genes associated with the rod phototransduction pathway between the three groups of organoids, we observed that several rod pathway components which were upregulated in patient organoids compared to control organoids were potentially salvaged following PR3 treatment (**Figure 6D**). EnrichGO analysis of significantly downregulated genes in visual perception/phototransduction pathways confirmed the specific activity of PR3 in retina (**Figure 6E**). While analysis for significantly upregulated genes showed changes in cilium organization, axoneme assembly, ER to Golgi transport and glycosylation, all of which are critically required for outer segment formation and protein trafficking (**Figure 6F, Supplementary Figure 5A-B**) suggestive of rod photoreceptor maturation recovery. Thus, the data presented strongly suggests that PR3 could potentially rescue rod photoreceptor homeostasis in *RHO*-CNV patients.

**Figure 6:**
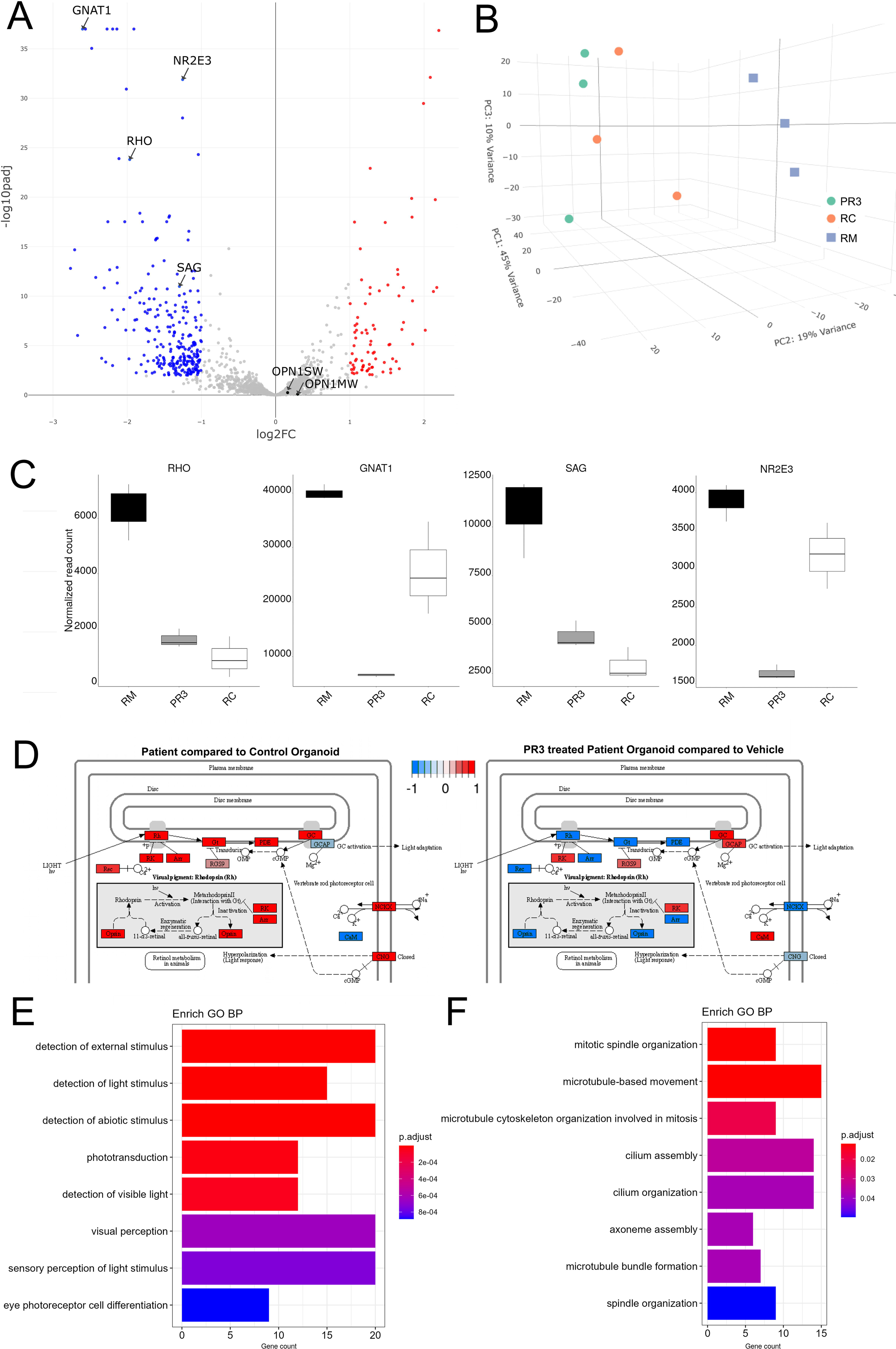
RNA sequencing analysis of *RHO*-CNV organoids following PR3 treatment. **(A)** Volcano plot showing significant differentially expressed genes following 1 week of 0.25µM PR3 treatment in D300+ patient organoids compared to vehicle treated with cutoffs at p<0.01 and ∼1log2FC. **(B)** 3D principal component analysis (PCA) plot showing the tightly clustered independent biological replicates from the control (RC) organoids, vehicle treated patient (RM) organoids and PR3 treated patient organoids (PR3). PR3 treated patient organoids were spatially closer to control (RC) compared to patient (RM) organoids. **(C)** Normalized read count plots showing relative expression of *RHO, GNAT1, SAG* and *NR2E3* in the three conditions. **(D)** KEGG analysis of differentially expressed genes showing dysregulation of key phototransduction pathway comparing RC with RM organoids and the recovery following PR3 treatment in RM organoids. Down- and up-regulated genes are indicated in blue and red respectively. **(E, F)** Box plots showing EnrichGO analysis of differentially expressed genes that are either downregulated (**E**) or upregulated (**F**) by comparing PR3 treated to vehicle treated RM organoids. N=3 independent experiments and 12-15 organoids per experiment. Log2FC=Log2 Fold change.

## Discussion

To our knowledge, there have been no previous reports of RP-associated with multiple copies of wild-type *RHO* in humans. However, retinal degeneration phenotypes have been reported in mice bearing extra copies of *Rho*. A 10-30% increase in rhodopsin expression in mice caused gradual degeneration, while a 3-5-fold increase resulted in severe and rapid photoreceptor loss (Olsson et al. 1992). More recent studies show that the retinal degeneration phenotype correlates closely to the amount of *RHO* overexpression (Tan et al. 2001; Wen et al. 2009). The current study reports the clinical retinal phenotype of a patient with *RHO*-CNV, and photoreceptor abnormalities in iPSC-derived retinal organoids from the patient. Using patient derived organoids provides unprecedented access to human patient specific material to better understand specific disease processes. These studies, also have a caveat to slow disease development due to the time involved in retinal maturation in organoid critical for pathological processes to kick-in. Another shortcoming of the current study is the lack of comparison of the organoid phenotype to an isogenic line. While the creation of isogenic lines and comparing them with disease organoid models is an ideal approach, carrying out large deletions by CRISPR-cas9 such as those required to fix the duplicated 188 kb and 48 kb inverted triplicated region in our *RHO*-CNV patient line is challenging effort. Large CRISPR edits generally have very low-efficiency (Eleveld et al. 2021; Canver et al. 2014). Thus, our study utilized an unaffected first-degree relative as a control.

In this patient, a few surrounding genes are duplicated and partially triplicated as well. Of these, IFT122 is particularly interesting as it has been shown to be part of the ciliary transport. IFT122 has been shown to cause recessive phenotype in dogs and in complete knockout zebrafish model but dominant or overexpression has not been shown to have a phenotype. We did a thorough survey through literature and through BluePrint Genetics database. We did not find any human retinal degeneration cases with variants in IFT122. Interestingly, recessive biallelic IFT122 mutation can cause cranioectodermal dysplasia (Sensenbrenner syndrome). However, none of these patients exhibited retinal dystrophy(Alazami et al. 2014). Among the other genes, HIF1OO is an epigenetic modifier gene, MBD4 is involved in DNA repair while PLXND1 is expressed in endothelial cells and none of these have been implicated in photoreceptor development or disease.

Our data strongly supports the notion that the RP phenotype in the patient is likely due to RHO accumulation and mislocalization in the patient’s photoreceptors. Based on studies in animal models, *RHO* mutations could lead to death of rod photoreceptors by several mechanisms, including: (a) overwhelming the transport machinery, thereby mislocalizing and driving cell dysfunction and death, as observed with Q344R/P/ter *RHO* mutations in mice(C. H. Sung et al. 1994); (b) reducing the supply of phototransduction deactivating proteins for chromophore-attached RHO, leading to constitutive activation and degeneration, as proposed in some forms of *RHO* mutations such as G90D and T94I(Dizhoor et al. 2008); (c) reducing RHO glycosylation in the Golgi leading to RHO instability and photoreceptor degeneration, as in T17M mutation(Murray et al. 2015), or by (d) RHO misfolding, driving ER stress/unfolded protein response (UPR)(Olsson et al. 1992; Chiang et al. 2015). Based on the RNAseq data of our current study, it is likely that RHO overexpression overloads the rod ER significantly reducing the efficiency of RHO glycosylation. Previous studies have shown that RHO undergoes N-linked glycosylation at two distinct sites, Asn-2 and Asn-15. This glycosylation is believed to be critical for protein folding through interactions with chaperones as well as for the transport of rhodopsin to the outer segments. Interestingly, studies in *Xenopus* suggest that this glycosylation is also responsible for nascent disc assembly(Murray, Fliesler, and AlLUbaidi 2009). While, we had initially hypothesized that RHO-CNV could lead to photoreceptor dysfunction due to ER stress/UPR dysfunction, we did not detect any such changes.

Mislocalization of RHO to the soma was the most striking phenotype discerned in our patient organoids. While there were some breakthroughs of RHO in putative OS, most of the expression especially in mature organoids (>D250) was stuck in the soma. Similar trafficking defects have been documented in post-mortem patient eye samples. RHO protein was found to be localized in the cell soma as well as synaptic processes and neurite extensions in RHO adRP patient samples as well as other RP patients (Li, Kljavin, and Milam 1995). This was also observed in transgenic pigs expressing human mutant RHO (Pro347Leu) (Li et al. 1998) as well as Xenopus expressing Q350ter (Tam et al. 2006). A remarkable feature of adRP is the late-stage retinal disease manifesting slow progressive degeneration with initial OS disruption prior to photoreceptor death. In our results, the *RHO*-CNV patient-matched retinal organoid model rigorously mimicked the clinically degenerative OS and intact photoreceptor phenotype. Interestingly the mislocalized RHO was not degraded over extended culture time of 300-days. Transgenic Rho mutant mice such as P23H mice have demonstrated the degradation of mislocalized RHO at 3 months of age (Price et al. 2011). These findings suggest that the death of the photoreceptors and vision loss in *RHO*-CNV patient is conversely different from patients with other adRP mutations including the P23H mutations. Hence it is essential to understand the underlying mechanism of rod photoreceptor death with similar mislocalized RHO cellular phenotype in patient-specific P23H and *RHO*-CNV retinal organoid models. Proof-of-principle studies utilizing PR3 have been established to alleviate the retinal pathology. Hence further studies comparing the P23H and *RHO*- CNV retinal organoid models could serve as a valuable cellular platform to evaluate the efficacy of potential genome editing and small molecule therapeutic strategies.

There is an increasing interest in gene augmentation therapeutic strategies for retinal degeneration patients with biallelic mutations in *RPE65*, since the approval of Voretigene neparvovec. Although visual outcomes in most treated patients have been very encouraging with significant improvement in visual field and light sensitivity(Maguire et al. 2021), there have been some recent reports of progressive pericentral atrophy(Gange et al. 2022). One potential cause of retinal pigmented epithelium (RPE) atrophy following *RPE65* gene augmentation could be overexpression of *RPE65* in the AAV-*RPE65* infected cells, due to the utilization of strong promoters such as CAG. Furthermore, the current preclinical approaches to target adRP due to *RHO* are primarily based on gene therapies, with three competing approaches; one to knockdown the mutant *RHO* by replacing with a wild-type *RHO*; second to merely overexpress wild-type RHO to overcome the mutant protein effect (Massengill and Lewin 2021), and third to overexpress NR2E3, the upstream regulator of *RHO* (S. Li et al. 2021). Overexpression of wild-type RHO approach could have unintended adverse consequences on photoreceptor survival if the relationship between RHO expression and rod survival is not clearly understood. Our current study on *RHO*-CNV associated with adRP raises the awareness of toxicities and complications that might arise from either of the gene augmentation therapy approaches.

Nuclear receptors are ideal therapeutic targets because their activities can be readily induced or repressed with small molecules. This allows for fine tuning of the biological functions of the receptor to alter disease pathogenesis. A few drugs targeting these receptors are already in the clinic (Dhiman, Bolt, and White 2018). It is interesting that targeting a reduction in wild-type RHO expression has also been proposed as a pan-RP therapy. Several groups have explored reduction in the expression of RHO as even small decreases can slow RP progression (Lewin et al. 1998). The photoregulin group of compounds, identified through a screen to target the *NR2E3* orphan nuclear receptor, were shown to repress rhodopsin expression with minor effects on cone opsins in mice (Nakamura et al. 2016). Photoregulins are suggested to act by enhancing the ability of NR2E3 to complex with NRL and CRX. PR3 was shown to reduce Rhodopsin expression in wild-type mice but not in rd7 mice bearing Nr2e3 mutations. Additionally, upon delivery to mice by intra-peritoneal injection, PR3 has been found to be safe and effective in mitigating retinal degeneration, and improving visual function in the P23H rhodopsin mouse model (Nakamura et al. 2017), suggesting photoregulins are attractive preclinical candidates to treat humans with *RHO*-related adRP. In our current study, PR3 had a robust effect on rhodopsin expression with no effects on cone opsins and could be a very useful therapeutic for patients with CNV. However in the case of other adRP *RHO* mutations, careful individualized dose management of the drug will likely be required to achieve adequate reduction of mutant rhodopsin expression in cases of adRP such that it no longer causes a toxic dominant negative effect while still allowing adequate wild-type RHO expression to allow for normal rod function as complete loss of rhodopsin can cause recessive RP.

In conclusion, we have established a disease-in-a-dish model for *RHO*-CNV associated adRP, and a potential therapeutic option for managing the devastating outcomes of the disorder.

## Conclusion

This is the first reported study of characterizing the *RHO*-CNV, a novel cause of adRP. This case of CNV has expanded the profile of highly mutated *RHO* by correlating the clinical behavior to disease modeling using retinal organoids. We have used the organoid model for targeted drug testing with PR3, small molecule inhibitor of *NR2E3*. Taken together, the results of this study demonstrate that *RHO* expression requires precise control for rod photoreceptor functioning and maintenance.

## Methods

### Clinical and Molecular Diagnosis

Both the proband (Male patient, 60-70 year-old) and his unaffected daughter (30-35-year-old) provided written informed consent, which was approved by the UCSF Institutional Review Board (IRB # 18-26409) and adhered to the tenets set forth in the Declaration of Helsinki. The participants gave consent to participate in the current study and have the results of this research work published. The proband (unidentified Lab ID # RM) and his asymptomatic daughter (unidentified Lab ID # RC) were examined at the University of California San Francisco (UCSF, CA, USA) including the pedigree data collection, clinical examination, and genetic diagnosis. Retinal examination included visual acuity testing, fundus photography, spectral domain optical coherence tomography (SD-OCT), full-field electroretinogram testing (ERG). For genetic diagnosis, peripheral blood (PB) samples collected from the proband and the unaffected daughter of the proband were screened by targeted Next Generation Sequencing (NGS) at Blueprint Genetic, a College of American Pathologists–and Clinical Laboratory Improvement Amendments–certified laboratory. The details of the methodology are available in our previous publication(Tuupanen et al. 2022). In brief, the sequencing was conducted using a retinal dystrophy NGS panel consisting of 266 genes, which was derived from an in-house tailored whole exome sequencing assay. The sequence reads were mapped to the human reference genome (GRCh37/hg19). CNV analysis was conducted using CNVkit (Talevich et al. 2016). Following a manual review of the identified RHO-CNV, discordant read-pairs suggesting a duplication-inverted triplication-duplication event was detected(Carvalho et al. 2011).

### Generating iPSC lines

Peripheral blood samples collected from the human subjects (unidentified Lab ID # RC and RM) were reprogrammed into iPSCs as per the schematic outlined in **Supplementary Figure 2A**. Blood samples were processed by density-gradient centrifugation using Ficoll-paque™ PLUS (17-1440-02, GE Healthcare Biosciences, Sweden) to isolate the peripheral blood mononuclear cells (PBMNCs) (**Supplementary Figure 2B)**. A small fraction of ∼1-2 x 10^6^ PBMNCs was cultured in 1-well of a 12-well suspension plate with peripheral blood mononuclear cells expansion medium (**Supplementary Table 1**) by changing half media every day. Date of seeding the PBMNCs in culture was designated as Day (D) −4. Four days later, ∼0.2-0.4 x 10^6^ PBMNCs were reprogrammed using CytoTune™-iPS 2.0 Sendai Reprogramming Kit (A16517, Thermo Fisher Scientific, USA), at 2.5:2.5:1.5 (KOS:c-myc: Klf4) multiplicity of infection. 24h post-transfection on D1, a full media change was done. Two days post transfection on D3, cells were plated onto 1-2 wells of a Matrigel (354234, Corning, USA, diluted as 1 µg/mL in DMEM/F12)-coated 6-well plate with reprogramming medium (**Supplementary Table 1**). Half media change was done every other day from D3 until D6. For all the suspension cultures, medium change was done by centrifugation at 200x *g* for 10 minutes and resuspending the cell pellet with the respective medium and culturing back into the plates. On D8, cells were transitioned to iPSC medium (**Supplementary Table 1**) with further media changes done on every other day. About 14-21 days post transfection, colonies with typical characteristics of iPSC morphology were manually picked. Pure, compact, and tightly packed iPSC clones were lifted using dissociating solution (**Supplementary Table 1**) for further expansion, characterization (**Supplementary Figure 2C-G)**, and retinal organoid differentiation.

### Retinal Organoid Differentiation

Retinal organoids were differentiated (three to six independent differentiation from each clones) using three different clones from each iPSC lines *via* the three step embryoid body (EB) approach following our previously published protocols and as per the schematic depicted in **Figure 3A**(Bachu et al. 2022; Arthur et al. 2022). Briefly, iPSC colonies were lifted and cultured in 6-well suspension plate. Small iPSC colonies self-aggregated as EBs within 24h were gradually transitioned to complete Neural Induction Medium (NIM) (**Supplementary Table 1**) from D0 to D3. On D6, EBs were treated with 1.5 nM BMP4 (120-05ET, PeproTech, USA). By D7, the EBs were transferred onto a Matrigel-coated plate to facilitate the adherence of EBs to the plate with NIM and 1.5 nM BMP4. Brief exposure of BMP4 in the differentiation process included the treatment of EBs with 1.5 nM (from D6-D8) to 0.75 nM (D9-D11) and 0.375 nM (D12-14). On D15, BMP4 was completely removed and EBs were fed with just NIM. From D16 through D30, the adherent EBs were fed Retinal Differentiation Medium (RDM) (**Supplementary Table 1**) at the intervals of every 2 days. Regions of the plate displaying clear, shiny borders indicative of retina-like morphology were manually lifted using a sterile P100 pipette tip. Lifted neural retina was transferred to a 6-well suspension plates and allowed to self-acquire the 3D organoid configuration within the next 2 days. From D31, the developing retinal organoids were fed 3D-RDM medium (**Supplementary Table 1**) along with 1 µM All-trans retinoic acid (R2625, Millipore Sigma) every 3days until D120. From D120 onwards, organoids were fed with just 3D-RDM medium without All-trans retinoic acid every 3 days until the completion of the study. Organoids were periodically assessed for morphological characteristics using phase contrast microscopy (Olympus IX70) at regular intervals. Various time-points of rod photoreceptor differentiation and maturation in the retinal organoids were utilized for phenotypic characterization and drug assessments (**Supplementary Figure 3B**).

### Immunohistochemistry and Imaging

Retinal organoids and confluent colonies of iPSCs were fixed in 4% paraformaldehyde (PFA) (157-8, Electron Microscopy Sciences, USA) in 1X PBS for 20 minutes. Organoids were cryopreserved in 15% through 30% sucrose (made in 1X PBS), and frozen in 2:1 mixture (20% sucrose: OCT, Sakura, USA). 10 µm sections of retinal organoids were collected on Super frost® Plus microscopic slides (12-550-15, Fisher Scientific, USA) using Leica CM3050 S cryostat. Immunohistochemistry was done as described previously. Organoid sections were pap-pen and PFA-fixed iPSC clones were permeabilized at room temperature for 15 mins with 0.1% Triton® X-100 (0694-1L, VWR Life Science, USA) in 10% Normal Donkey Serum (NDS, S30-100ML, EMD Millipore Corp, USA; made in 1X PBS). Following to that, sections were incubated for 1 hour in 10% NDS. Primary antibodies (**Supplementary Table 3A**) diluted in 10% NDS were added to the slides and incubated overnight (12-18 h) at 4°C. The slides were washed thrice in 1X PBS at 5 mins intervals and further incubated for 1 hour at room temperature with fluorescently conjugated secondary antibodies (**Supplementary Table 3B**) diluted at 1:250 in 10% NDS. Cell nuclei were counter stained with DAPI (1ug/mL, Roche, USA) for 10 minutes. The slides were then washed thrice in 1X PBS at 5 mins interval and cover slipped using Fluoromount G (Electron Microscopy Sciences, USA). TUNEL analysis was carried out per manufacturer’s protocol (Sigma-Aldrich). Images were acquired using ZEN software on LSM700 confocal microscope (Zeiss, Inc.) and processed by Image J (NIH, USA).

### RNA extraction and Quantitative Real-Time PCR (qRT-PCR)

Total RNA was extracted from retinal organoids using RNeasy® Mini Kit (74104, Qiagen, USA) as per the manufacturer’s instructions. The quality and purity of the extracted RNA was assessed by NanoDrop One (Thermo Fisher Scientific, USA). cDNA was synthesized using iScript cDNA Synthesis Kit (1708891, Bio-Rad, USA). Real-time qRT-PCR was performed on CFX96 system (Bio-Rad, USA) using iTaq™ Universal SYBR® Green Supermix (64047467, Bio-Rad, USA). Primer sequences used for qRT-PCR is listed in **Supplementary Table 4**. The amplification reactions set were: 95°C for 30 s, 40 cycles at 95°C for 5 s, 60°C for 25 s, 95°C for 5 s and final extension at 95°C for 5 s. The Ct values of the target genes were first normalized to the endogenous control β-actin. The corrected Ct values were then utilized to compare and validate the control and patient retinal organoids. Log_2_ fold change (log2FC) in gene expression of all targets were then calculated.

### Bulk RNA Sequencing

The quality control, RNA library preparations and sequencing reactions of the extracted RNA from retinal organoids were performed at Novogene Corporation Inc (CA, USA). FASTQ files received from Novogene following QC were quantified using Salmon package (V 1.9.0) by pseudo-aligning against homo sapiens hg19 genomic assembly in Galaxy(Afgan et al. 2016). Low-counts (<10) were filtered out and batch correction was carried out using Combat package in DEBrowser (v1.24.1) R package (Kucukural et al. 2019). Differential expression analysis was carried out using DESeq2 in DEbrowser. Genes with an adjusted p-value < 0.05 were assigned as differentially expressed. Gene ontology enrichment analysis and KEGG pathway analysis of differentially expressed genes was carried out in DEBrowser and Beavr (Perampalam and Dick 2020)packages. Differential expression analysis and pathway enrichment data have been uploaded as supplementary files (**Table 2**). The data is uploaded to NCBI GEO database (GSE245545).

### Transmission Electron Microscopy (TEM)

Retinal organoids (300-days-old) were washed in 1X PBS three times at 10 mins intervals and fixed with Karnovsky’s fixative (4% paraformaldehyde/2% glutaraldehyde in 0.1 M PBS, pH 7.4) overnight at 4°C. For TEM, organoids were washed in 1X PBS and stained in 1% osmium tetroxide (19180, Electron Microscopy Sciences, USA; made in distilled water) for 1 hour at room temperature followed by staining with 2% uranyl acetate (22400, Electron Microscopy Sciences, USA; made in distilled water). After three washes in distilled water, organoids were then dehydrated in a graded series of ice-cold ethanol (50%, 70%, 95% and 100%) for 20 mins each. Organoids were incubated in propylene oxide (00235-1, Polysciences Inc, USA) twice for 5 mins, followed by a 5-mins incubation in 1:1 mix of propylene oxide: Epon 812 (13940, Electron Microscopy Sciences, USA). Organoids were allowed to infiltrate in Epon 812 overnight at room temperature. Next day, the organoids were embedded in PELCO silicone rubber molds (105, Ted Pella Inc, USA) using Epon 812, and polymerized for 48 hours at 60°C. Ultrathin (70 nm) sections were collected and imaged using a Philips Tecnai 10 electron microscope at the VA Medical Center, San Francisco, USA.

### Western Blotting

Retinal organoids were collected and washed in cold 1X PBS twice at 5 mins intervals. Protein was extracted by homogenizing the organoids using hand-held pestle (1415-5390, USA Scientific) in ice-cold RIPA buffer (**See supplementary Table 2A**) with 1% complete™ Protease inhibitor cocktail (11697498001, Roche Diagnostics, GmbH). Lysates were incubated at 4°C for 10 minutes and then centrifuged at 14,000 *xg* for 15 minutes at 4°C. Supernatant was collected and the protein concentration was measured using Pierce™ BCA Protein Assay Kit (23227, Thermo Scientific, USA) as per the manufacturer’s instructions. Equal amounts (20 µg) of protein lysates were mixed with 4x Laemmli sample buffer (161-0747, Bio-Rad, USA) and 10% dithiothreitol (DTT, R0861, Thermo Scientific, USA). Samples were resolved on a precast gel (12% Mini-Protean TGX Gels, 456-1044, Bio-Rad, USA) for 20 mins at 70 V until the loading dye entered the resolving layer, then increased to 150 V for 40-60 mins or until the run was completed (**See supplementary Table 2A**). After electrophoresis, the proteins were transferred onto an Immobilon^®^-FL PVDF membrane (IPFL20200, Merck Millipore, USA) ice-cold transfer buffer (**See supplementary Table 2A**) at 100V for 90 minutes. The blots were blocked with blocking solution for 30 minutes. The blots were incubated overnight at 4°C with primary antibodies diluted in blocking solution (**See supplementary Table 3A for primary antibody details**). Thereafter, blots were washed thrice with TBS-T buffer (10 mins interval each) and incubated for 1 hour at room temperature with host-specific secondary antibodies diluted in blocking solution (**See supplementary Table 3B for secondary antibody details**). The blots were washed another three times with TBS-T buffer (10 mins interval each) and visualized using LiCor Odyssey XF scanner. For quantification of protein expression, the background was subtracted for each band and normalized to internal control band using Image J software. Statistical calculations were performed using multiple technical and five independent biological replicates.

### PR3 treatment on Retinal Organoids

Patient-specific *RHO*-CNV retinal organoids (∼300-days-old) were treated with small-drug like molecule Photoregulin3 (PR3, SML2299-5MG, Sigma, 5 mM stock made in DMSO) supplemented in 3D-RDM for 1 week by changing medium every other day. Three different concentrations of PR3 utilized in this study were 0.1, 0.25, and 0.5 µM. DMSO was used as a vehicle control. One-week, post-PR3 treatment, retinal organoids were collected and assessed by qRT-PCR, bulk RNA sequencing and imaging experiments.

### Statistical Analysis

All the data were obtained from three to six independent retinal differentiation experiments. The quantified data values were provided as mean ± SEM. The intergroup differences for all the analysis were determined with the GraphPad Prism v9, using a two-tailed student’s t-test or ANOVA. Significant differences were indicated by p values listed in the figures. For imaging studies, at least 3 sections were averaged to account for regional variability in iPSC reprograming and retinal organoid differentiation.

## Supporting information

Suppl-RNAseq _data

## Data Availability

All data produced in the present study will be available upon reasonable request to the authors.

## Acknowledgments

We are grateful to the recruited human subjects and families included in this study. We thank members of the Lamba lab for helpful discussions and suggestions. The research presented here is supported by the R01 EY032197 and CIRM DISC0-14449 to D.A.L, Foundation Fighting Blindness research grant to D.A.L, U24 EY029891 to D.A.L and J.L.D, Postdoctoral fellowship from All May See Foundation and Bright Focus Foundation macular degeneration research to S.K, UCSF Vision Core NIH/NEI P30 EY002162, and unrestricted grant from Research to Prevent Blindness, New York, NY to Department of Ophthalmology at UCSF. We deeply appreciate the help of Yien-Ming Kuo at the UCSF Vision Core and Ivy Hsieh at VA Medical Center for processing the electron microscopy samples. We would also like to thank Suling Wang for drawing all the illustrations included in this paper based on the descriptions given by the authors. Additionally, we would like to acknowledge the contributions of The Foundation Fighting Blindness My Retina Tracker® Genetic Testing Program for identifying the *RHO*-CNV in the patient. We deeply appreciate the services given by Blueprint Genetics who undertook the genetic analysis and particularly Sari Tuupanen who resolved the complex genetic rearrangement within the *RHO* locus.

---

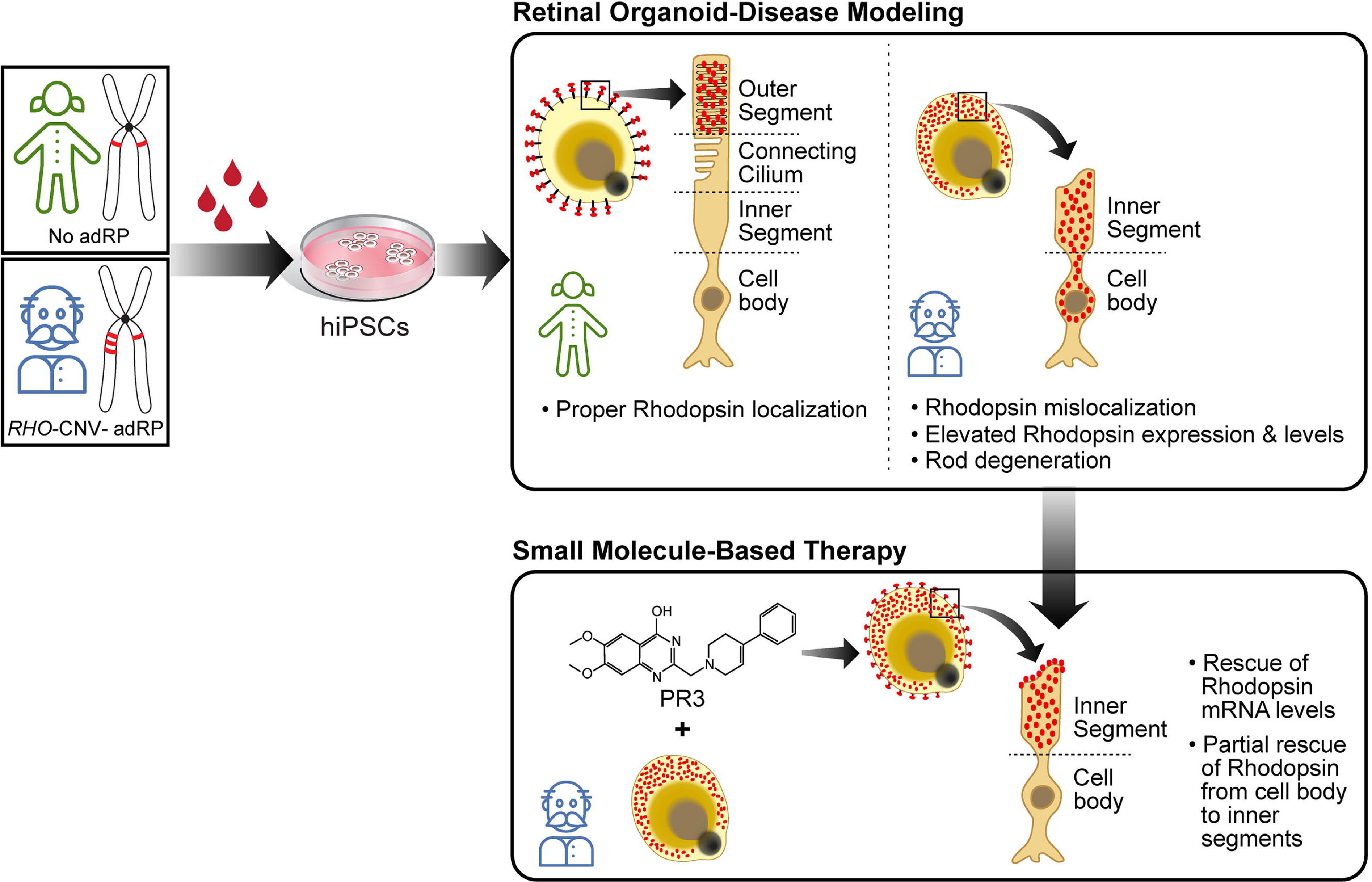

## References

Afgan, Enis, Dannon Baker, Marius van den Beek, Daniel Blankenberg, Dave Bouvier, Martin Čech, John Chilton, et al. 2016. “The Galaxy Platform for Accessible, Reproducible and Collaborative Biomedical Analyses: 2016 Update.” Nucleic Acids Research 44 (W1): W3–10. 10.1093/nar/gkw343.

Alazami, Anas M, Mohammed Zain Seidahmed, Fatema Alzahrani, Adam O Mohammed, and Fowzan S Alkuraya. 2014. “Novel IFT122 Mutation Associated with Impaired Ciliogenesis and Cranioectodermal Dysplasia.” Molecular Genetics & Genomic Medicine 2 (2): 103–6. 10.1002/mgg3.44.

Arthur, Peggy, Sangeetha Kandoi, Li Sun, Anil Kalvala, Shallu Kutlehria, Santanu Bhattacharya, Tanmay Kulkarni, et al. 2022. “Biophysical, Molecular and Proteomic Profiling of Human Retinal Organoid-Derived Exosomes.” Pharmaceutical Research, August. 10.1007/s11095-022-03350-7.

Athanasiou, Dimitra, Monica Aguila, James Bellingham, Wenwen Li, Caroline McCulley, Philip J Reeves, and Michael E Cheetham. 2018. “The Molecular and Cellular Basis of Rhodopsin Retinitis Pigmentosa Reveals Potential Strategies for Therapy.” Progress in Retinal and Eye Research 62 (January): 1–23. 10.1016/j.preteyeres.2017.10.002.

Bachu, Vismaya S, Sangeetha Kandoi, Ko Uoon Park, Michael L Kaufman, Michael Schwanke, Deepak A Lamba, and Joseph A Brzezinski. 2022. “An Enhancer Located in a Pde6c Intron Drives Transient Expression in the Cone Photoreceptors of Developing Mouse and Human Retinas.” Developmental Biology 488 (August): 131–50. 10.1016/j.ydbio.2022.05.012.

Bumsted O’Brien, Keely M, Hong Cheng, Yibin Jiang, Dorothea Schulte, Anand Swaroop, and Anita E Hendrickson. 2004. “Expression of Photoreceptor-Specific Nuclear Receptor NR2E3 in Rod Photoreceptors of Fetal Human Retina.” Investigative Ophthalmology & Visual Science 45 (8): 2807–12. 10.1167/iovs.03-1317.

Carvalho, Claudia M B, Melissa B Ramocki, Davut Pehlivan, Luis M Franco, Claudia Gonzaga-Jauregui, Ping Fang, Alanna McCall, et al. 2011. “Inverted Genomic Segments and Complex Triplication Rearrangements Are Mediated by Inverted Repeats in the Human Genome.” Nature Genetics 43 (11): 1074–81. 10.1038/ng.944.

Cheng, Hong, Hemant Khanna, Edwin C T Oh, David Hicks, Kenneth P Mitton, and Anand Swaroop. 2004. “Photoreceptor-Specific Nuclear Receptor NR2E3 Functions as a Transcriptional Activator in Rod Photoreceptors.” Human Molecular Genetics 13 (15): 1563–75. 10.1093/hmg/ddh173.

Chiang, Wei-Chieh, Heike Kroeger, Sanae Sakami, Carissa Messah, Douglas Yasumura, Michael T Matthes, Judith A Coppinger, Krzysztof Palczewski, Matthew M LaVail, and Jonathan H Lin. 2015. “Robust Endoplasmic Reticulum-Associated Degradation of Rhodopsin Precedes Retinal Degeneration.” Molecular Neurobiology 52 (1): 679–95. 10.1007/s12035-014-8881-8.

Chirco, Kathleen R, Shereen Chew, Anthony T Moore, Jacque L Duncan, and Deepak A Lamba. 2021. “Allele-Specific Gene Editing to Rescue Dominant CRX-Associated Leber Congenital Amaurosis (LCA7) Phenotypes in a Retinal Organoid Model.” Stem Cell Reports 16 (November): 1–13.

Chizzolini, Marzio, Alessandro Galan, Elisabeth Milan, Adolfo Sebastiani, Ciro Costagliola, and Francesco Parmeggiani. 2011. “Good Epidemiologic Practice in Retinitis Pigmentosa: From Phenotyping to Biobanking.” Current Genomics 12 (4): 260–66. 10.2174/138920211795860071.

Dhiman, Vineet K, Michael J Bolt, and Kevin P White. 2018. “Nuclear Receptors in Cancer - Uncovering New and Evolving Roles through Genomic Analysis.” Nature Reviews. Genetics 19 (3): 160–74. 10.1038/nrg.2017.102.

Dizhoor, Alexander M, Michael L Woodruff, Elena V Olshevskaya, Marianne C Cilluffo, M Carter Cornwall, Paul A Sieving, and Gordon L Fain. 2008. “Night Blindness and the Mechanism of Constitutive Signaling of Mutant G90D Rhodopsin.” The Journal of Neuroscience 28 (45): 11662–72. 10.1523/JNEUROSCI.4006-08.2008.

Dryja, T P, T L McGee, E Reichel, L B Hahn, G S Cowley, D W Yandell, M A Sandberg, and E L Berson. 1990. “A Point Mutation of the Rhodopsin Gene in One Form of Retinitis Pigmentosa.” Nature 343 (6256): 364–66. 10.1038/343364a0.

Dulla, Kalyan, Ralph Slijkerman, Hester C van Diepen, Silvia Albert, Margo Dona, Wouter Beumer, Janne J Turunen, et al. 2021. “Antisense Oligonucleotide-Based Treatment of Retinitis Pigmentosa Caused by USH2A Exon 13 Mutations.” Molecular Therapy 29 (8): 2441–55. 10.1016/j.ymthe.2021.04.024.

Duncan, Jacque L, Karmen Trzupek, Joan Fisher, Leilla Kenney, Sari Tuupanen, Miika Mehine, Stephen P Daiger, and Brian Mansfield. 2019. “Multiple Copies of Rhodopsin as a Novel Cause of Autosomal Dominant Retinitis Pigmentosa.” Investigative Ophthalmology & Visual Science 60 (9): 2943–2943.

Giacalone, Joseph C, Jeaneen L Andorf, Qihong Zhang, Erin R Burnight, Dalyz Ochoa, Austin J Reutzel, Malia M Collins, et al. 2019. “Development of a Molecularly Stable Gene Therapy Vector for the Treatment of RPGR-Associated X-Linked Retinitis Pigmentosa.” Human Gene Therapy 30 (8): 967–74. 10.1089/hum.2018.244.

Hirji, Sitara H. 2023. “Clinical Evaluation of Patients with Retinitis Pigmentosa.” Methods in Molecular Biology 2560: 31–39. 10.1007/978-1-0716-2651-1_3.

Kobayashi, M, S Takezawa, K Hara, R T Yu, Y Umesono, K Agata, M Taniwaki, K Yasuda, and K Umesono. 1999. “Identification of a Photoreceptor Cell-Specific Nuclear Receptor.” Proceedings of the National Academy of Sciences of the United States of America 96 (9): 4814–19. 10.1073/pnas.96.9.4814.

Kruczek, Kamil, Zepeng Qu, James Gentry, Benjamin R Fadl, Linn Gieser, Suja Hiriyanna, Zachary Batz, et al. 2021. “Gene Therapy of Dominant CRX-Leber Congenital Amaurosis Using Patient Stem Cell-Derived Retinal Organoids.” Stem Cell Reports 16 (2): 252–63. 10.1016/j.stemcr.2020.12.018.

Kucukural, Alper, Onur Yukselen, Deniz M Ozata, Melissa J Moore, and Manuel Garber. 2019. “DEBrowser: Interactive Differential Expression Analysis and Visualization Tool for Count Data.” BMC Genomics 20 (1): 6. 10.1186/s12864-018-5362-x.

Lewin, A S, K A Drenser, W W Hauswirth, S Nishikawa, D Yasumura, J G Flannery, and M M LaVail. 1998. “Ribozyme Rescue of Photoreceptor Cells in a Transgenic Rat Model of Autosomal Dominant Retinitis Pigmentosa.” Nature Medicine 4 (8): 967–71. 10.1038/nm0898-967.

Liu, Lisa, Lei Yu, Zhichao Li, Wujiao Li, and WeiRen Huang. 2021. “Patient-Derived Organoid (PDO) Platforms to Facilitate Clinical Decision Making.” Journal of Translational Medicine 19 (1): 40. 10.1186/s12967-020-02677-2.

Li, Z Y, I J Kljavin, and A H Milam. 1995. “Rod Photoreceptor Neurite Sprouting in Retinitis Pigmentosa.” The Journal of Neuroscience 15 (8): 5429–38. 10.1523/JNEUROSCI.15-08-05429.1995.

Li, Z Y, F Wong, J H Chang, D E Possin, Y Hao, R M Petters, and A H Milam. 1998. “Rhodopsin Transgenic Pigs as a Model for Human Retinitis Pigmentosa.” Investigative Ophthalmology & Visual Science 39 (5): 808–19.

Meng, Da, Sara D Ragi, and Stephen H Tsang. 2020. “Therapy in Rhodopsin-Mediated Autosomal Dominant Retinitis Pigmentosa.” Molecular Therapy 28 (10): 2139–49. 10.1016/j.ymthe.2020.08.012.

Mitton, K P, P K Swain, S Chen, S Xu, D J Zack, and A Swaroop. 2000. “The Leucine Zipper of NRL Interacts with the CRX Homeodomain. A Possible Mechanism of Transcriptional Synergy in Rhodopsin Regulation.” The Journal of Biological Chemistry 275 (38): 29794–99. 10.1074/jbc.M003658200.

Moore, Spencer M, Dorota Skowronska-Krawczyk, and Daniel L Chao. 2020. “Targeting of the NRL Pathway as a Therapeutic Strategy to Treat Retinitis Pigmentosa.” Journal of Clinical Medicine 9 (7). 10.3390/jcm9072224.

Murray, Anne R, Steven J Fliesler, and Muayyad R Al-Ubaidi. 2009. “Rhodopsin: The Functional Significance of Asn-Linked Glycosylation and Other Post-Translational Modifications.” Ophthalmic Genetics 30 (3): 109–20. 10.1080/13816810902962405.

Murray, Anne R, Linda Vuong, Daniel Brobst, Steven J Fliesler, Neal S Peachey, Marina S Gorbatyuk, Muna I Naash, and Muayyad R Al-Ubaidi. 2015. “Glycosylation of Rhodopsin Is Necessary for Its Stability and Incorporation into Photoreceptor Outer Segment Discs.” Human Molecular Genetics 24 (10): 2709–23. 10.1093/hmg/ddv031.

Nakamura, Paul A, Andy A Shimchuk, Shibing Tang, Zhizhi Wang, Kole DeGolier, Sheng Ding, and Thomas A Reh. 2017. “Small Molecule Photoregulin3 Prevents Retinal Degeneration in the RhoP23H Mouse Model of Retinitis Pigmentosa.” ELife 6 (November). 10.7554/eLife.30577.

Nakamura, Paul A, Shibing Tang, Andy A Shimchuk, Sheng Ding, and Thomas A Reh. 2016. “Potential of Small Molecule-Mediated Reprogramming of Rod Photoreceptors to Treat Retinitis Pigmentosa.” Investigative Ophthalmology & Visual Science 57 (14): 6407–15. 10.1167/iovs.16-20177.

Newton, Fay, and Roly Megaw. 2020. “Mechanisms of Photoreceptor Death in Retinitis Pigmentosa.” Genes 11 (10). 10.3390/genes11101120.

O’Neal, Teri B., and Euil E. Luther. 2023. “Retinitis Pigmentosa.” In StatPearls. Treasure Island (FL): StatPearls Publishing.

Olsson, J E, J W Gordon, B S Pawlyk, D Roof, A Hayes, R S Molday, S Mukai, G S Cowley, E L Berson, and T P Dryja. 1992. “Transgenic Mice with a Rhodopsin Mutation (Pro23His): A Mouse Model of Autosomal Dominant Retinitis Pigmentosa.” Neuron 9 (5): 815–30. 10.1016/0896-6273(92)90236-7.

Parfitt, David A, Amelia Lane, Conor Ramsden, Katarina Jovanovic, Peter J Coffey, Alison J Hardcastle, and Michael E Cheetham. 2016. “Using Induced Pluripotent Stem Cells to Understand Retinal Ciliopathy Disease Mechanisms and Develop Therapies.” Biochemical Society Transactions 44 (5): 1245–51. 10.1042/BST20160156.

Perampalam, Pirunthan, and Frederick A Dick. 2020. “BEAVR: A Browser-Based Tool for the Exploration and Visualization of RNA-Seq Data.” BMC Bioinformatics 21 (1): 221. 10.1186/s12859-020-03549-8.

Price, Brandee A, Ivette M Sandoval, Fung Chan, David L Simons, Samuel M Wu, Theodore G Wensel, and John H Wilson. 2011. “Mislocalization and Degradation of Human P23H-Rhodopsin-GFP in a Knockin Mouse Model of Retinitis Pigmentosa.” Investigative Ophthalmology & Visual Science 52 (13): 9728–36. 10.1167/iovs.11-8654.

“RetNet: Summaries.” n.d. Accessed June 26, 2023. https://web.sph.uth.edu/RetNet/sum-dis.htm.

Sung, Ching-Hwa, and Andrew W. Tai. 1999. “Rhodopsin Trafficking and Its Role in Retinal Dystrophies.” In, 195:215–67. International Review of Cytology. Elsevier. 10.1016/S0074-7696(08)62706-0.

Sung, C H, C Makino, D Baylor, and J Nathans. 1994. “A Rhodopsin Gene Mutation Responsible for Autosomal Dominant Retinitis Pigmentosa Results in a Protein That Is Defective in Localization to the Photoreceptor Outer Segment.” The Journal of Neuroscience 14 (10): 5818–33. 10.1523/JNEUROSCI.14-10-05818.1994.

Talevich, Eric, A Hunter Shain, Thomas Botton, and Boris C Bastian. 2016. “CNVkit: Genome-Wide Copy Number Detection and Visualization from Targeted DNA Sequencing.” PLoS Computational Biology 12 (4): e1004873. 10.1371/journal.pcbi.1004873.

Tam, Beatrice M, and Orson L Moritz. 2009. “The Role of Rhodopsin Glycosylation in Protein Folding, Trafficking, and Light-Sensitive Retinal Degeneration.” The Journal of Neuroscience 29 (48): 15145–54. 10.1523/JNEUROSCI.4259-09.2009.

Tam, Beatrice M, Guifu Xie, Daniel D Oprian, and Orson L Moritz. 2006. “Mislocalized Rhodopsin Does Not Require Activation to Cause Retinal Degeneration and Neurite Outgrowth in Xenopus Laevis.” The Journal of Neuroscience 26 (1): 203–9. 10.1523/JNEUROSCI.3849-05.2006.

Tucker, Budd A, Robert F Mullins, Luan M Streb, Kristin Anfinson, Mari E Eyestone, Emily Kaalberg, Megan J Riker, Arlene V Drack, Terry A Braun, and Edwin M Stone. 2013. “Patient-Specific IPSC-Derived Photoreceptor Precursor Cells as a Means to Investigate Retinitis Pigmentosa.” ELife 2 (August): e00824. 10.7554/eLife.00824.

Tuupanen, Sari, Kimberly Gall, Johanna Sistonen, Inka Saarinen, Kati Kämpjärvi, Kirsty Wells, Katja Merkkiniemi, et al. 2022. “Prevalence of RPGR-Mediated Retinal Dystrophy in an Unselected Cohort of Over 5000 Patients.” Translational Vision Science & Technology 11 (1): 6. 10.1167/tvst.11.1.6.

Volland, Stefanie, Julian Esteve-Rudd, Juyea Hoo, Claudine Yee, and David S Williams. 2015. “A Comparison of Some Organizational Characteristics of the Mouse Central Retina and the Human Macula.” Plos One 10 (4): e0125631. 10.1371/journal.pone.0125631.

Yamamoto, Haruka, Tetsuo Kon, Yoshihiro Omori, and Takahisa Furukawa. 2020. “Functional and Evolutionary Diversification of Otx2 and Crx in Vertebrate Retinal Photoreceptor and Bipolar Cell Development.” Cell Reports 30 (3): 658–671.e5. 10.1016/j.celrep.2019.12.072.

Zhao, Linjie, Shengtao Zhou, and Jan-Åke Gustafsson. 2019. “Nuclear Receptors: Recent Drug Discovery for Cancer Therapies.” Endocrine Reviews 40 (5): 1207–49. 10.1210/er.2018-00222.

