## Supplementary figures and images for "Disease modeling and pharmacological rescue of autosomal dominant Retinitis Pigmentosa associated with *RHO* copy number variation"

### Suppl. Figures

#### Suppl. Figure 1

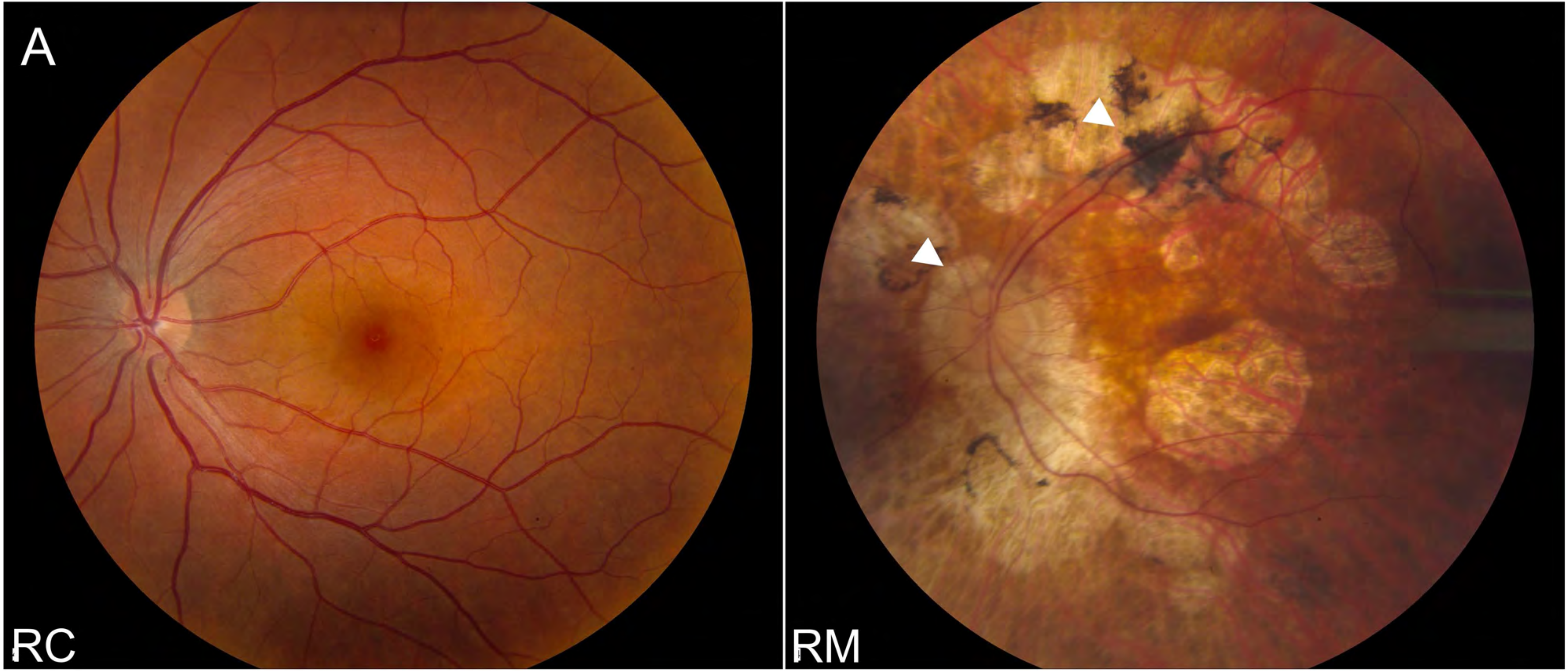

#### Suppl. Figure 2

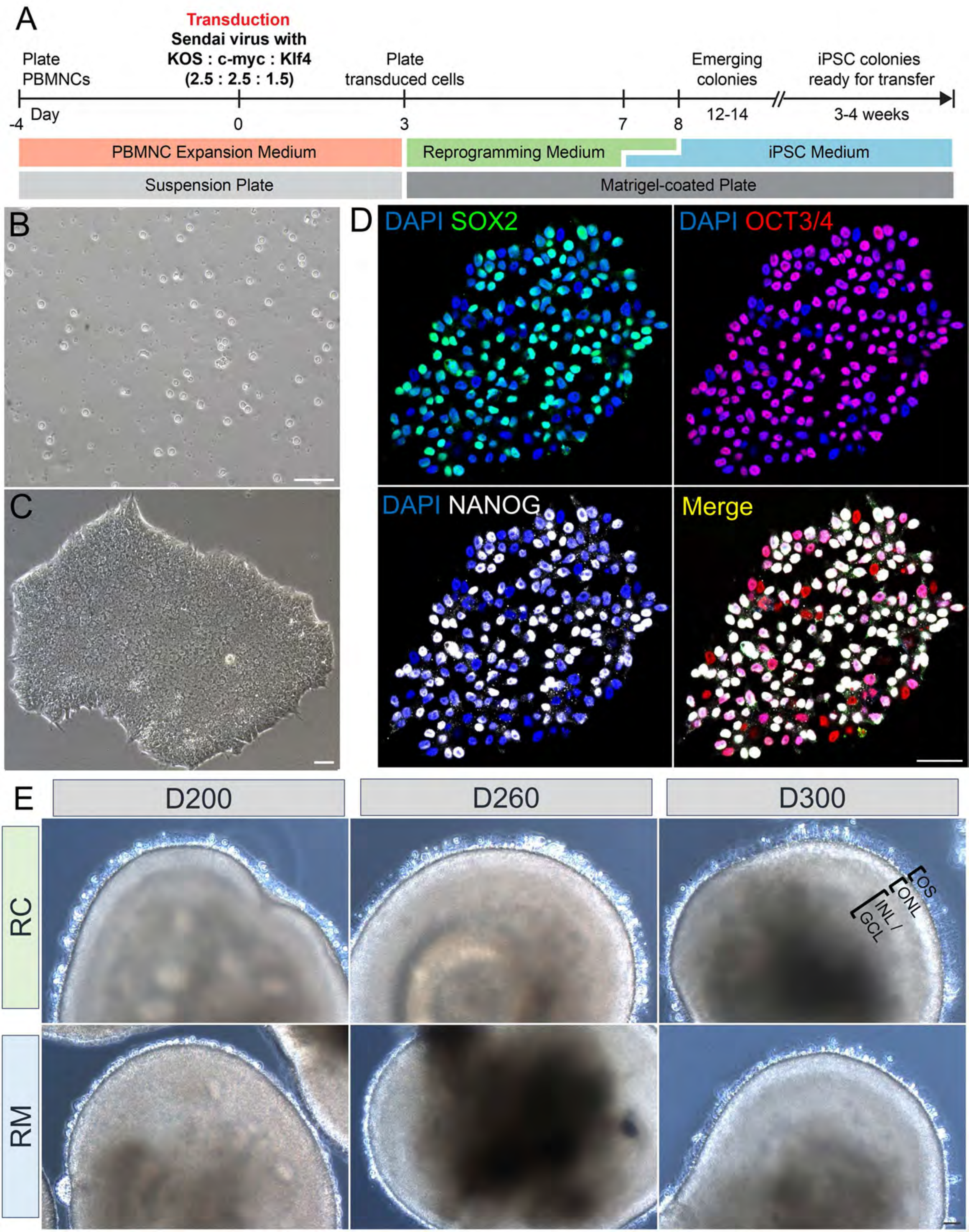

Suppl.
Figure
3

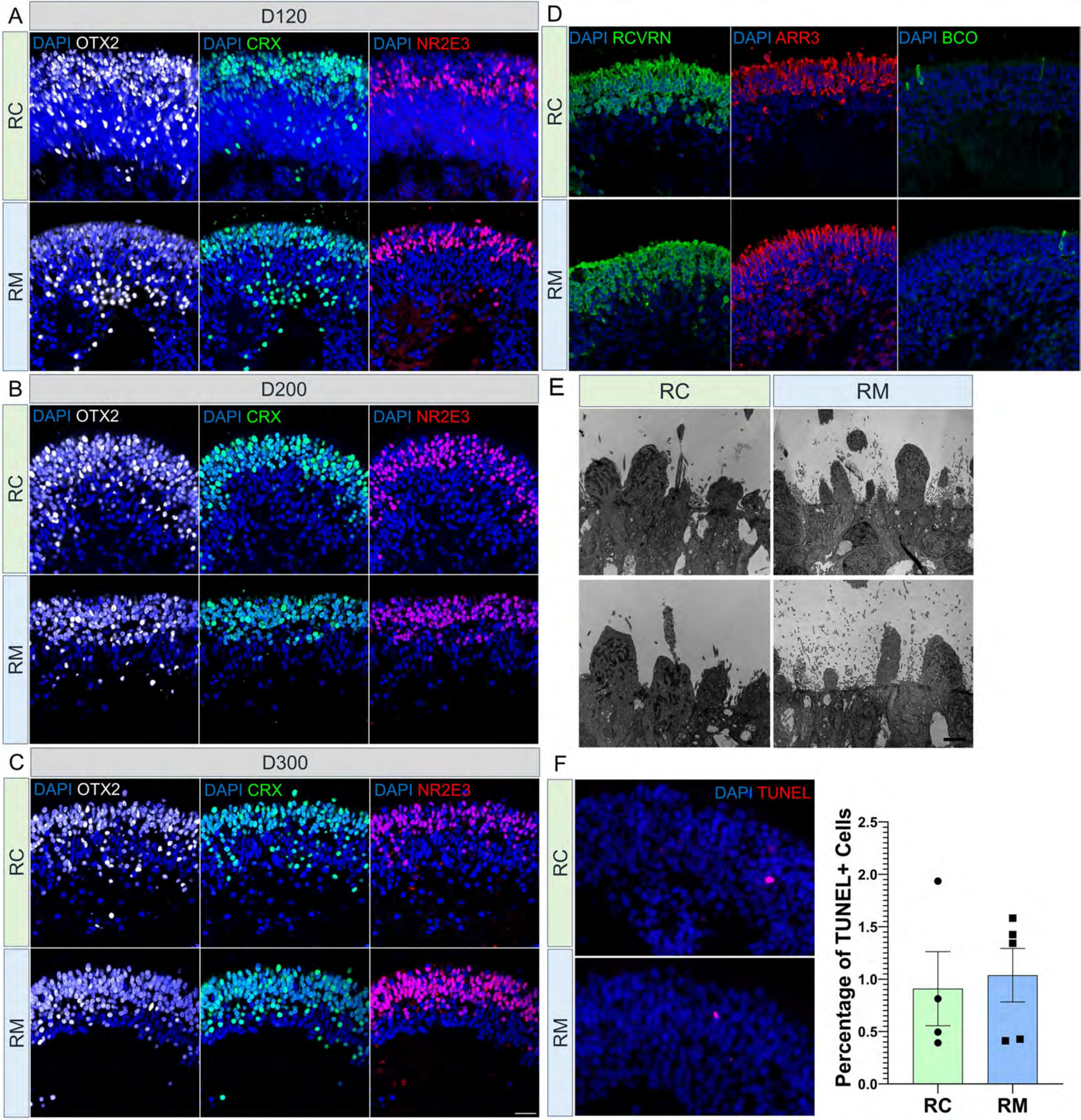

#### Suppl. Figure 4

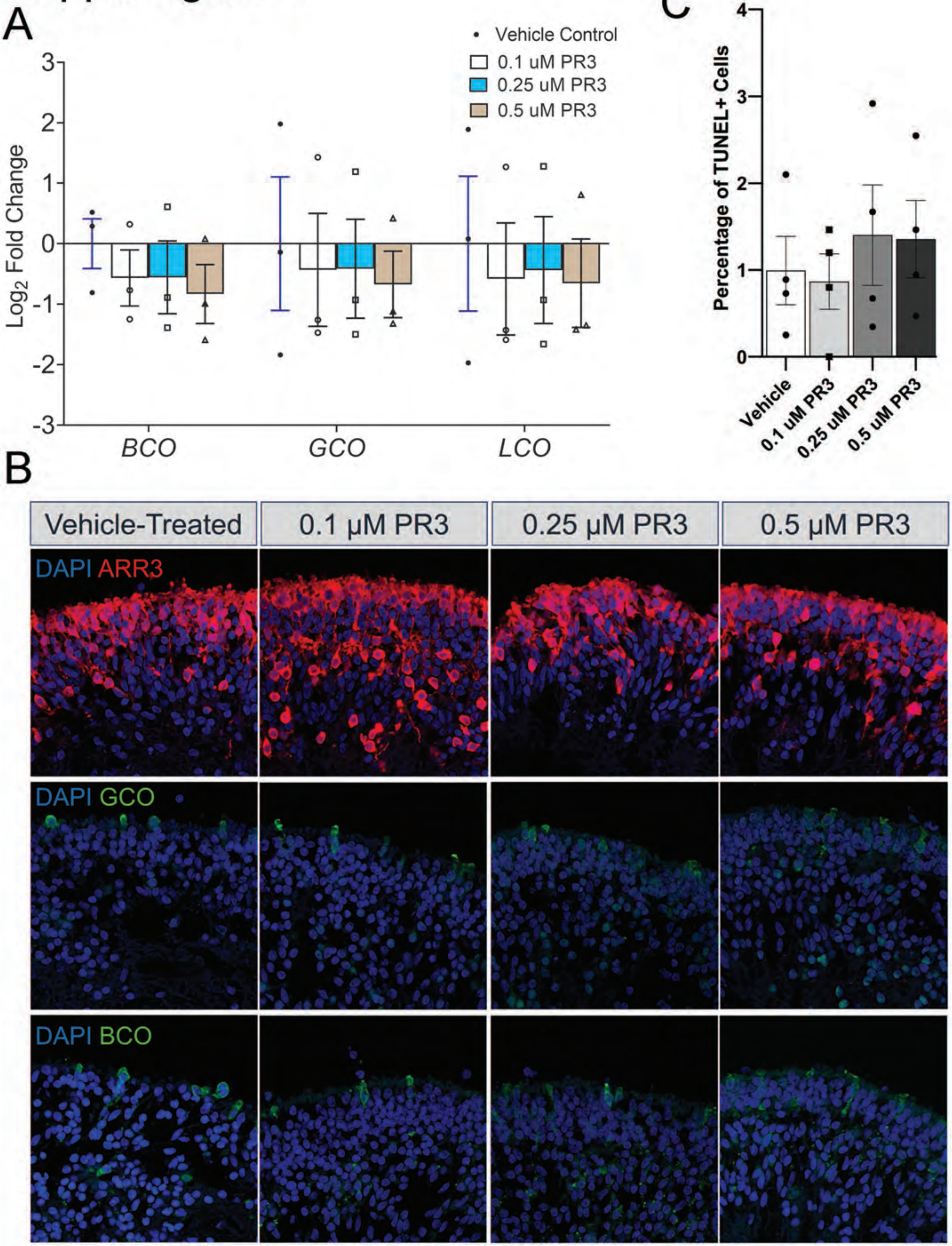

#### Suppl. Figure 5

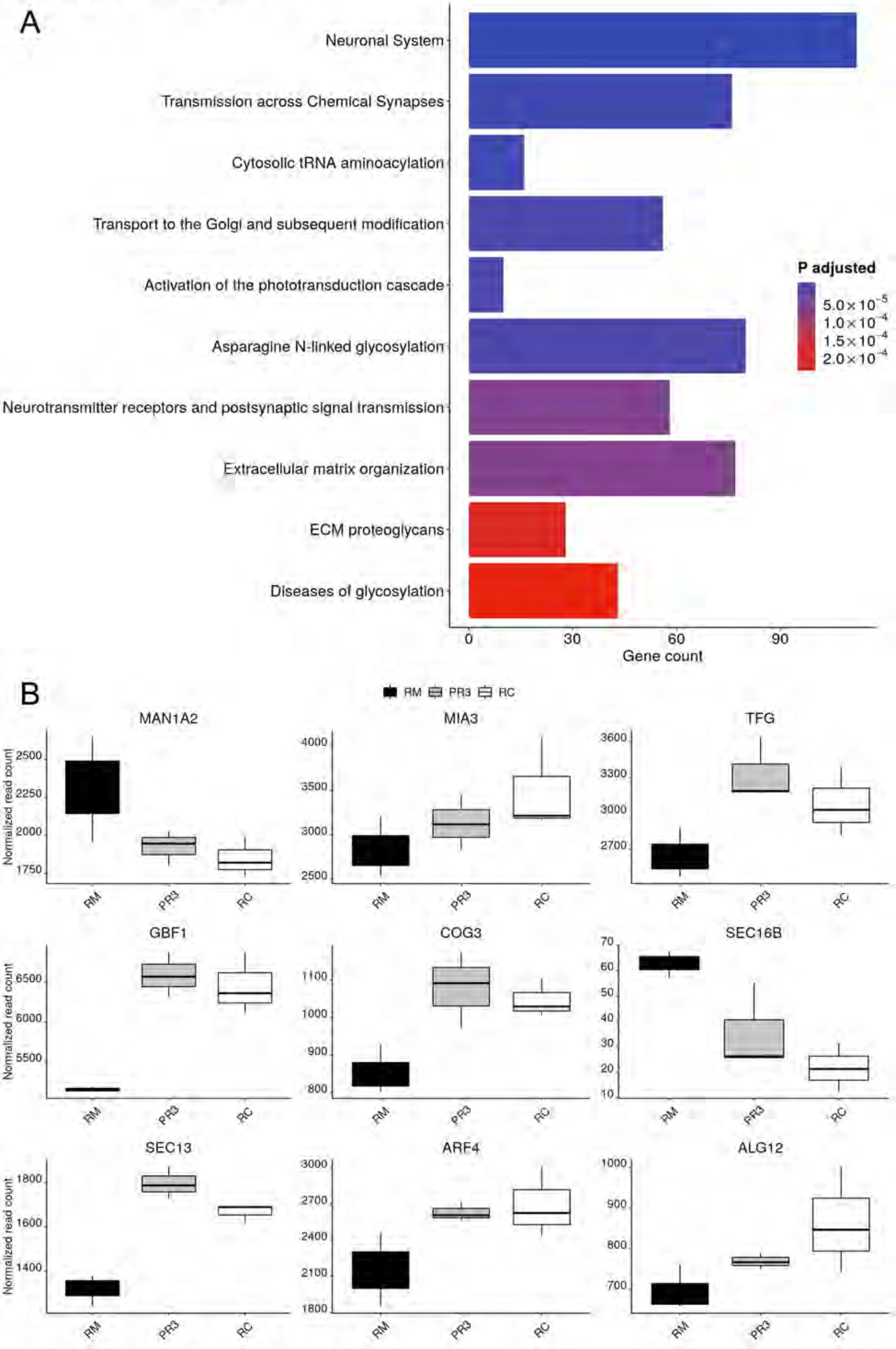
