## Supplementary material for "Disease modeling and pharmacological rescue of autosomal dominant Retinitis Pigmentosa associated with *RHO* copy number variation": Suppl. Tables

**Supplementary Table 1**. Media Formulations.

**PBMNC Expansion Medium**

| **Component** | **Brand** | **Reference #** | **Stock Concentration** | **Final Concentration** |
| --- | --- | --- | --- | --- |
| StemPro^®^-34 SFM (1X) | Gibco | 10640-019 | N/A | Remainder |
| GlutaMax™-I (100X) | Gibco | 35050-061 | N/A | 1X |
| Recombinant Human SCF | PeproTech, USA | 300-07 | 50 µg/mL | 100 ng/uL |
| Recombinant Human Flt3-Ligand | PeproTech, USA | 300-19 | 50 µg/mL | 100 ng/uL |
| Recombinant Human IL-3 | PeproTech, USA | 200-03 | 50 µg/mL | 20 ng/uL |
| Recombinant Human IL-6 | PeproTech, USA | 200-06 | 20 µg/mL | 20 ng/uL |

**Abbreviations:** Flt3, IL, interleukin; N/A, not applicable; PBMNC, peripheral blood mononuclear cells; SCF, stem cell factor.

**Reprogramming Medium**

| **Component** | **Brand** | **Reference #** | **Final Concentration** |
| --- | --- | --- | --- |
| StemPro^®^-34 SFM (1X) | Gibco | 10640-019 | N/A |
| GlutaMax™-I (100X) | Gibco | 35050-061 | 1X |

**Abbreviations:** N/A, not applicable.

**iPSC Medium**

| **Component** | **Brand** | **Reference #** | **Final Concentration** |
| --- | --- | --- | --- |
| mTeSTR™ Plus Medium | Stemcell Technologies | 100-0276 | N/A |
| GlutaMax™-I (100X) | Gibco | 35050-061 | 1X |
| Antibiotic/Antimycotic Solution (100X) | Cytiva | SV30079.01 | 1X |

**Abbreviations:** N/A, not applicable.

**Dissociating Solution**

| **Component** | **Brand** | **Reference #** | **Final Concentration** |
| --- | --- | --- | --- |
| DPBS, 1X | Corning | 21-031-CV | N/A |
| EDTA, 0.5 M, pH 8.0 +/- 0.1 | Corning | 46-034-Cl | 0.1% |

**Abbreviations:** N/A, not applicable.

**NIM**

| **Component** | **Brand** | **Reference #** | **Stock Concentration** | **Final Concentration** |
| --- | --- | --- | --- | --- |
| DMEM/F12 (1:1) (1X) | Gibco | 11330-032 | N/A | N/A |
| GlutaMax™-I (100X) | Gibco | 35050-061 | N/A | 1X |
| MEM Nonessential Amino Acids (100X) Solution | Corning | 25-025-Cl | N/A | 1X |
| Heparin sodium salt from porcine intestinal mucosa | Sigma-Aldrich | H3149-100KU | 2 mg/mL | 2 µg/mL |
| Antibiotic/Antimycotic Solution (100X) | Cytiva | SV30079.01 | N/A | 1X |
| FILTER | | | | |
| N-2 Supplement (100X) | Gibco | 17502-001 | 100X | 1X |

**Abbreviations:** N/A, not applicable; NIM, Neural induction medium.

**RDM**

| **Component** | **Brand** | **Reference #** | **Stock Concentration** | **Final Concentration** |
| --- | --- | --- | --- | --- |
| DMEM/High Glucose | Cytiva | SH30022.02 | N/A | 1:1 |
| DME/F-12 1:1 (1X) | Cytiva | SH30023.02 | N/A |  |
| GlutaMax™-I (100X) | Gibco | 35050-061 | 100X | 1X |
| MEM Nonessential Amino Acids (100X) Solution | Corning | 25-025-Cl | 100X | 1X |
| Antibiotic/Antimycotic Solution (100X) | Cytiva | SV30079.01 | 100X | 1X |
| FILTER | | | | |
| B-27® Supplement (50X) | Gibco | 17502-001 | 50X | 1X |

**Abbreviations:** N/A, not applicable; RDM, Retinal differentiation medium.

**3D-RDM**

| **Component** | **Brand** | **Reference #** | **Stock Concentration** | **Final Concentration** |
| --- | --- | --- | --- | --- |
| DMEM/High Glucose | Cytiva | SH30022.02 | N/A | 1:1 |
| DME/F-12 1:1 (1X) | Cytiva | SH30023.02 | N/A |  |
| GlutaMax™-I (100X) | Gibco | 35050-061 | 100X | 1X |
| Antibiotic/Antimycotic Solution (100X) | Cytiva | SV30079.01 | 100X | 1X |
| Taurine | Sigma Life Science | T0625-10G | 100 mM | 200 µM |
| Fetal Bovine Serum, Premium | Atlanta Biologicals | S11150 | N/A | 5% |
| FILTER | | | | |
| B-27® Supplement (50X) | Gibco | 17502-001 | 50X | 1X |
| CD Lipid Concentrate | Gibco | 11905-031 | N/A | 0.1% |

**Abbreviations:** N/A, not applicable; 3D-RDM, 3 Dimensional-Retinal differentiation media.

**Supplementary Table 2**. Western Blotting Reagents.

**RIPA Buffer**

| **Component** | **Brand** | **Reference #** | **Stock Concentration** | **Final Concentration** |
| --- | --- | --- | --- | --- |
| Tris/HCL, 1M solution, pH-7.5 Ultrapure MB grade | Affymetrix | 22639-500ML | 1M | 50mM |
| Sodium Chloride | Fisher Scientific | BP358-10 | 5M | 150mM |
| Sodium dodecyl sulfate | Invitrogen | AM9820 | 20% | 0.1% (Volume/Volume) |
| Deoxycholic acid, sodium salt | Calbiochem | 264101-100GM | NA | 0.5% (Weight/Volume) |
| Triton-X-100 | Sigma Life Science | T9284-100ML | 20% | 1% (Volume/Volume) |
| pH-7.5 | | | | |

**Abbreviations:** N/A, not applicable; RIPA, Radioimmunoprecipitation assay buffer.

**SDS-PAGE Running Buffer**

| **Component** | **Brand** | **Reference #** | **Final Concentration** |
| --- | --- | --- | --- |
| 10X Tris/Glycine/SDS Buffer | Bio-Rad | 1610732 | 100 mL |
| Distilled Water | N/A | N/A | 900 mL |
| Total | | | 1000 mL |

**Transfer Buffer**

| **Component** | **Brand** | **Reference #** | **Final Concentration** |
| --- | --- | --- | --- |
| 10X Tris/Glycine Buffer | Bio-Rad | 1610734 | 100 mL |
| Methanol | Honeywell | 179957-2.5L | 100 mL |
| Distilled Water | N/A | N/A | 800 mL |
| Total | | | 1000 mL |

**Blocking Solution**

| **Component** | **Brand** | **Reference #** | **Final Concentration** |
| --- | --- | --- | --- |
| Intercept blocking solution | Licor, USA | 927-65001 | 50% |
| 10X TBS buffer | Bio-Rad | 1706435 | 50% of 1X TBS  (Dilute to 10X TBS to 1X in distilled water before use) |

**TBS-T Buffer**

| **Component** | **Brand** | **Reference #** | **Final Concentration** |
| --- | --- | --- | --- |
| 10X TBS buffer | Bio-Rad | 1706435 | 100 mL of 1X TBS  (Dilute to 10X TBS to 1X in distilled water before use) |
| Distilled Water | N/A | N/A | 899 mL |
| Tween 20 | Sigma Life Science | P9416-100ML | 1 mL |
| Total | | | 1000 mL |

**Supplementary Table 3A**. Primary Antibody List.

| **Protein Name** | **Host** | **Dilution** | **Source** | **Reference #** | **Purpose** |
| --- | --- | --- | --- | --- | --- |
| β-ACTIN | Rabbit | 1:4000 | Abcam | ab8227 | Western Blotting |
| CRX | Rabbit | 1:400 | Abcam | ab140603 | Immunohistochemistry |
| NANOG (D73G4) | Rabbit | 1:400 | Cell Signaling | 4903S | Immunohistochemistry |
| NRL | Goat | 1:400 | R&D Systems | AF2945 | Immunohistochemistry |
| NR2E3/PNR | Mouse | 1:400 | R&D Systems | PP-H7223-00 | Immunohistochemistry |
| OCT3/4 (C-10) | Mouse | 1:100 | Santa Cruz Biotechnology, Inc. | sc-5279 | Immunohistochemistry |
| OTX2 | Goat | 1:400 | R&D Systems | BAF1979 | Immunohistochemistry |
| RHO (1D4) | Mouse | 1:500 | Santa Cruz Biotechnology, Inc. | sc-57432 | Western Blotting |
| RHO (4D2) | Mouse | 1:500 | EMD Millipore | MABN15 | Immunohistochemistry |
| SAG (E-3) | Mouse | 1:200  1:500 | Santa Cruz Biotechnology, Inc. | sc-166383 | Immunohistochemistry (1:200)  Western Blotting (1:500) |
| ARR3 | Mouse | 1:250 | EMD Millipore | MABN2636 | Immunohistochemistry |
| BCO | Rabbit | 1:100 | EMD Millipore | AB5407 | Immunohistochemistry |
| GCO | Rabbit | 1:250 | EMD Millipore | AB5405 | Immunohistochemistry |
| RCVRN | Rabbit | 1:500 | EMD Millipore | AB5585 | Immunohistochemistry |
| SOX2 | Goat | 1:400 | R&D Systems | AF2018 | Immunohistochemistry |

**Abbreviations:** β-ACTIN, Beta actin; CRX, Cone-rod homeobox protein; NRL, Nuclear retina leucine zipper; NR2E3/PNR, Nuclear receptor subfamily 2 group E member 3/Photoreceptor-Specific Nuclear Receptor; OCT3/4, Octamer-binding protein 3/4; OTX2, Orthodenticle homolog 2; RHO, Rhodopsin/Opsin-2; SAG, Retinal S-antigen/rod photoreceptor arrestin/S-arrestin; ARR3, Cone arrestin; BCO, Blue cone opsin; GCO, Green cone opsin; RCVRN, Recoverin.

**Table 3B**. Secondary Antibody List.

| **Species** | **Target** | **Conjugate** | **Dilution** | **Source** | **Cat. No.** |
| --- | --- | --- | --- | --- | --- |
| Donkey | Anti-mouse | Alexa Fluor 488 | 1:250 | Invitrogen | A-21202 |
| Donkey | Anti-mouse | Alexa Fluor 555 | 1:250 | Invitrogen | A-31570 |
| Donkey | Anti-mouse | Alexa Fluor 647 | 1:250 | Invitrogen | A-31571 |
| Donkey | Anti-rabbit | Alexa Fluor 488 | 1:250 | Invitrogen | A-21206 |
| Donkey | Anti-rabbit | Alexa Fluor 555 | 1:250 | Invitrogen | A-31572 |
| Donkey | Anti-rabbit | Alexa Fluor 647 | 1:250 | Invitrogen | A-31573 |
| Donkey | Anti-Goat | Alexa Fluor 488 | 1:250 | Invitrogen | A-11055 |
| Donkey | Anti-Goat | Alexa Fluor 555 | 1:250 | Invitrogen | A-21432 |
| Donkey | Anti-Goat | Alexa Fluor 647 | 1:250 | Invitrogen | A-21447 |
| Goat | Anti-mouse | DyLight™ 800 4X PEG | 1:4000 | Invitrogen | SA535521 |
| Goat | Anti-rabbit | DyLight™ 680 | 1:4000 | Thermo Fisher Scientific | 35568 |

**Supplementary Table 4**. qRT-PCR Primer List.

| **Gene Symbol** | **Full Name** | **Forward primer (5’→3’)** | **Reverse primer (5’→3’)** |
| --- | --- | --- | --- |
| *ACTB* | Actin beta | GACCTGACTGACTACCTCAT | GTAGCACAGCTTCTCCTTAAT |
| *CRX* | Cone-Rod Homeobox | CAGCTAGGAGGTTACAGATTG | GTCACTCTTCCTGGCTTTAG |
| *GNAT1* | G Protein Subunit Alpha Transducin 1 | CTGAAAGAGGACGCTGAGAAG | TACCCGTCCTGGTGGATAAT |
| *IFT122* | Intraflagellar Transport 122 | TGCTGAGACCTACCTGAAGA | AGGATGCTTCTCACCCAAAG |
| *NR2E3* | Nuclear Receptor Subfamily 2 Group E Member 3 | TGATGTCACCAGCAATGACC | TCTTCCAGCAGGATCACCT |
| *NRL* | Neural Retina Leucine Zipper | ATGTGGATTGGACGACTTC | TTGGCGAGATTGTCTTGG |
| *OTX2* | Orthodenticle Homeobox 2 | CGAGAGGAGGTGGCACTGA | TGTTGTTGGCGGCACTTAGC |
| *PDE6B* | Phosphodiesterase 6B | AAGTGGGCTTCATCGACTTC | CGCCTTCCACTCTTTCCTATT |
| *RCVRN* | Recoverin | CACACACACATGCACACACACG | TGTGTGCAGGCCTTTCTCTTGG |
| *RHO* | Rhodopsin | TCATCATGGTCATCGCTTTC | CATGAAGATGGGACCGAAGT |
| *SAG* | S-Antigen Visual Arrestin | CACAGAGAAGACCGTGAAGAAG | CCACGGGCTTGACGTAATAA |
| *OPN1LW (LCO)* | Opsin 1, Long Wave Sensitive  (Long/Red cone opsin) | CACCTTCTTCGCATGCTTTG | TCGAAACTGCCGGTTCATAA |
| *OPN1MW (GCO)* | Opsin 1, Medium Wave Sensitive  (Green cone opsin) | TGGTCTCTGGCCATCATTTC | AGTACCTGCTCCAACCAAAG |
| *OPN1SW (BCO)* | Opsin 1, Short Wave Sensitive  (Blue cone opsin) | CTTCCGCTTCAGCTCCAA | GAACCGGCTCCAGCCA |
